# A Checklist for Assessing the Methodological Quality of Concurrent tES-fMRI Studies (ContES Checklist): A Consensus Study and Statement

**DOI:** 10.1101/2020.12.23.20248579

**Authors:** Hamed Ekhtiari, Peyman Ghobadi-Azbari, Axel Thielscher, Andrea Antal, Lucia M. Li, A Duke Shereen, Yuranny Cabral-Calderin, Daniel Keeser, Til Ole Bergmann, Asif Jamil, Ines R. Violante, Jorge Almeida, Marcus Meinzer, Hartwig R. Siebner, Adam J. Woods, Charlotte J. Stagg, Rany Abend, Daria Antonenko, Tibor Auer, Marc Bächinger, Chris Baeken, Helen C. Barron, Henry W. Chase, Jenny Crinion, Abhishek Datta, Matthew H. Davis, Mohsen Ebrahimi, Zeinab Esmaeilpour, Brian Falcone, Valentina Fiori, Iman Ghodratitoostani, Gadi Gilam, Roland H. Grabner, Joel D. Greenspan, Georg Groen, Gesa Hartwigsen, Tobias U. Hauser, Christoph S. Herrmann, Chi-Hung Juan, Bart Krekelberg, Stephanie Lefebvre, Sook-Lei Liew, Kristoffer H. Madsen, Rasoul Mahdavifar-Khayati, Nastaran Malmir, Paola Marangolo, Andrew K. Martin, Timothy J. Meeker, Hossein Mohaddes Ardabili, Marius Moisa, Davide Momi, Beni Mulyana, Alexander Opitz, Natasza Orlov, Patrick Ragert, Christian C. Ruff, Giulio Ruffini, Michaela Ruttorf, Arshiya Sangchooli, Klaus Schellhorn, Gottfried Schlaug, Bernhard Sehm, Ghazaleh Soleimani, Hosna Tavakoli, Benjamin Thompson, Dagmar Timmann, Aki Tsuchiyagaito, Martin Ulrich, Johannes Vosskuhl, Christiane A. Weinrich, Mehran Zare-Bidoky, Xiaochu Zhang, Benedikt Zoefel, Michael A. Nitsche, Marom Bikson

**Affiliations:** Laureate Institute for Brain Research, Tulsa, OK, USA; Department of Biomedical Engineering, Shahed University, Tehran, Iran; Iranian National Center for Addiction Studies (INCAS), Tehran University of Medical Sciences, Tehran, Iran; Danish Research Centre for Magnetic Resonance, Centre for Functional and Diagnostic Imaging and Research, Copenhagen University Hospital Amager and Hvidovre, Hvidovre, Denmark; Department of Health Technology, Technical University of Denmark, Kgs. Lyngby, Denmark; Department of Neurology, University Medical Center Goettingen, Goettingen, Germany; Computational, Cognitive and Clinical Imaging Lab, Division of Brain Sciences, Department of Medicine, Imperial College London, UK; UK DRI Centre for Care Research and Technology, Imperial College London, UK; Advanced Science Research Center, The Graduate Center, City University of New York, New York, NY, USA; Research Group Neural and Environmental Rhythms, Max Planck Institute for Empirical Aesthetics, Frankfurt, Germany; Department of Psychiatry and Psychotherapy, University Hospital LMU Munich, Munich, Germany; Department of Radiology, University Hospital LMU Munich, Munich, Germany; Neuroimaging Center (NIC), Focus Program Translational Neuroscience (FTN), Johannes Gutenberg University Medical Center, Langenbeckstr. 1, 55131, Mainz, Germany; Leibniz Institute for Resilience Research, Wallstraße 7, 55122, Mainz, Germany; Department of Neurology and Stroke and Hertie Institute for Clinical Brain Research, Eberhard Karls University of Tübingen, Tübingen, Germany; Department of Psychology and Neurosciences, Leibniz Research Centre for Working Environment and Human Factors, Dortmund, Germany; School of Psychology, Faculty of Health and Medical Sciences, University of Surrey, Guildford, UK; Proaction Lab, Faculty of Psychology and Educational Sciences, University of Coimbra, Portugal; CINEICC, Faculty of Psychology and Educational Sciences, University of Coimbra, Portugal; Centre for Clinical Research (UQCCR), The University of Queensland, Brisbane, Australia; Department of Neurology, University Medicine Greifswald, Greifswald, Germany; Department of Neurology, Copenhagen University Hospital Bispebjerg and Frederiksberg, Copenhagen, Denmark; Institute of Clinical Medicine, University of Copenhagen, Copenhagen, Denmark; Center for Cognitive Aging and Memory, McKnight Brain Institute, Department of Clinical and Health Psychology, University of Florida, Gainesville, FL, USA; Wellcome Centre for Integrative Neuroimaging, University of Oxford, FMRIB, John Radcliffe Hospital, Oxford, UK; Medical Research Council Brain Network Dynamics Unit, Nuffield Department of Clinical Neurosciences, University of Oxford, Mansfield Road, Oxford, UK; Section on Development and Affective Neuroscience, National Institute of Mental Health, Bethesda, MD, USA; Neural Control of Movement Lab, Department of Health Sciences and Technology, Zurich, Switzerland; Neuroscience Center Zurich, University of Zurich, Swiss Federal Institute of Technology Zurich, Zurich, Switzerland; Department of Psychiatry and Medical Psychology, University Hospital Ghent, Belgium; Department of Psychiatry, Vrije Universiteit Brussel, University Hospital Brussels, Belgium; Eindhoven University of Technology, Department of Electrical Engineering, the Netherlands; Department of Psychiatry, University of Pittsburgh School of Medicine, Pittsburgh, PA, USA; Institute of Cognitive Neuroscience, University College London, London, UK; Research and Development, Soterix Medical, New York, USA; The City College of the City University of New York, New York, USA; MRC Cognition and Brain Sciences Unit, University of Cambridge, Cambridge, UK; Department of Biomedical Engineering, The City College of New York of CUNY, New York, NY, USA; Northrop Grumman Company, Mission Systems, Falls Church, VA, United States; Department of Clinical and Behavioral Neurology, IRCCS Santa Lucia Foundation, Rome, Italy; Neurocognitive Engineering Laboratory (NEL), Center for Engineering Applied to Health, Institute of Mathematics and Computer Science (ICMC), University of Sao Paulo, Brazil; Systems Neuroscience and Pain Laboratory, Division of Pain Medicine, Department of Anesthesiology, Perioperative, and Pain Medicine, School of Medicine, Stanford University, Palo Alto, CA, USA; Educational Neuroscience, Institute of Psychology, University of Graz, Austria; Department of Neural and Pain Sciences, University of Maryland School of Dentistry, Baltimore, MD, USA; Department of Psychiatry, University of Ulm, Ulm, Germany; Lise Meitner Research Group Cognition and Plasticity, Max Planck Institute for Human Cognitive and Brain Sciences, Leipzig, Germany; Max Planck University College London Centre for Computational Psychiatry and Ageing Research, University College London, London, UK; Wellcome Centre for Human Neuroimaging, University College London, London, UK; Experimental Psychology Lab, Cluster of Excellence “Hearing4all”, European Medical School, University of Oldenburg, Oldenburg, Germany; Neuroimaging Unit, European Medical School, University of Oldenburg, Oldenburg, Germany; Research Centre Neurosensory Science, University of Oldenburg, Oldenburg, Germany; Institute of Cognitive Neuroscience, National Central University, Taoyuan, Taiwan; Cognitive Intelligence and Precision Healthcare Research Center, National Central University, Taoyuan, Taiwan; Center for Molecular and Behavioral Neuroscience, Rutgers University-Newark, Newark, New Jersey, USA; Translational Research Centre, University Hospital of Psychiatry, University of Bern, Bern, Switzerland; Chan Division of Occupational Science and Occupational Therapy, University of Southern California, Los Angeles, California, USA; USC Stevens Neuroimaging and Informatics Institute, Department of Neurology, Keck School of Medicine, University of Southern California, Los Angeles, California, USA; Department of Biomedical Engineering, University of Southern California, Los Angeles, California, USA; Division of Biokinesiology and Physical Therapy, University of Southern California, Los Angeles, California, USA; Department of Applied Mathematics and Computer Science, Technical University of Denmark, Kgs. Lyngby, Denmark; Department of Humanities Studies, University Federico II, Naples, Italy; Aphasia Research Lab, IRCCS Santa Lucia Foundation, Rome, Italy; Department of Psychology, University of Kent, Canterbury, UK; Department of Neurosurgery, Johns Hopkins University, Baltimore, Maryland; Psychiatry and Behavioral Sciences Research Center, Ibn-e-Sina Hospital, Faculty of Medicine, Mashhad University of Medical Sciences, Mashhad, Iran; Student Research Committee, Faculty of Medicine, Mashhad University of Medical Sciences, Mashhad, Iran; Zurich Center for Neuroeconomics, Department of Economics, University of Zurich, Zurich, Switzerland; Krembil Centre for Neuroinformatics, Centre for Addiction and Mental Health (CAMH), Toronto, Canada; Department of Biomedical Engineering, University of Minnesota, Minneapolis, MN, USA; Department of Psychosis Studies, Institute of Psychiatry, Psychology and Neuroscience, King’s College London, London, UK; Athinoula A. Martinos Center for Biomedical Imaging, Massachusetts General Hospital, Boston, MA, USA; Department of Radiology, Xuanwu Hospital, Capital Medical University, Beijing, China; Department of Psychology, Jagiellonian University, Cracow, Poland; Institute for General Kinesiology and Exercise Science, University of Leipzig, Leipzig, Germany; Department of Neurology, Max Planck Institute for Human Cognitive and Brain Sciences, Leipzig, Germany; Neuroelectrics Corporation, Cambridge, Massachusetts, MA, USA; Neuroelectrics Barcelona, Barcelona, Spain; Computer Assisted Clinical Medicine, Medical Faculty Mannheim, Heidelberg University, Mannheim, Germany; neuroConn GmbH, Ilmenau, Germany; Neuroimaging-Neuromodulation and Stroke Recovery Laboratories, Department of Neurology, Baystate-University of Massachusetts Medical School, and Department of Biomedical Engineering, Institute of Applied Life Sciences, UMass Amherst, MA, USA; Department of Biomedical Engineering, Amirkabir University of Technology, Tehran, Iran; Department of Cognitive Neuroscience, Institute for Cognitive Sciences Studies, Tehran, Iran; School of Optometry and Vision Science, University of Auckland, Auckland, New Zealand; School of Optometry and Vision Science, University of Waterloo, Ontario, Canada; Centre for Eye and Vision Research, Hong Kong, Hong Kong; Department of Neurology and Center for Translational Neuro- and Behavioral Sciences (C-TNBS), Essen University Hospital, University of Duisburg-Essen, Essen, Germany; Department of Cognitive Neurology, University Medical Center Goettingen, Goettingen, Germany; Shahid-Sadoughi University of Medical Sciences, Yazd, Iran; Department of Psychology, School of Humanities & Social Science, University of Science & Technology of China, Hefei, China; Centre de Recherche Cerveau et Cognition (CerCo), CNRS, Toulouse, France; Université Toulouse III Paul Sabatier, Toulouse, France; Department of Neurology, University Medical Hospital Bergmannsheil, Bochum, Germany

**Keywords:** Transcranial electrical stimulation (tES), Functional magnetic resonance imaging (fMRI), Concurrent tES fMRI, Consensus statement, ContES checklist

## Abstract

**Background:** Low intensity transcranial electrical stimulation (tES), including alternating or direct current stimulation (tACS or tDCS), applies weak electrical stimulation to modulate the activity of brain circuits. Integration of tES with concurrent functional magnetic resonance imaging (fMRI) allows for the mapping of neural activity during neuromodulation, supporting causal studies of both brain function and tES effects. Methodological aspects of tES-fMRI studies underpin the results, and reporting them in appropriate detail is required for reproducibility and interpretability. Despite the growing number of published reports, there are no consensus-based checklists for disclosing methodological details of concurrent tES-fMRI studies.

**Objective:** To develop a consensus-based checklist of reporting standards for concurrent tES-fMRI studies to support methodological rigor, transparency, and reproducibility (ContES Checklist).

**Methods:** A two-phase Delphi consensus process was conducted by a steering committee (SC) of 13 members and 49 expert panelists (EP) through the International Network of the tES-fMRI (INTF) Consortium. The process began with a circulation of a preliminary checklist of essential items and additional recommendations, developed by the SC based on a systematic review of 57 concurrent tES-fMRI studies. Contributors were then invited to suggest revisions or additions to the initial checklist. After the revision phase, contributors rated the importance of the 17 essential items and 42 additional recommendations in the final checklist. The state of methodological transparency within the 57 reviewed concurrent tES-fMRI studies was then assessed using the checklist.

**Results:** Experts refined the checklist through the revision and rating phases, leading to a checklist with three categories of essential items and additional recommendations: (1) technological factors, (2) safety and noise tests, and (3) methodological factors. The level of reporting of checklist items varied among the 57 concurrent tES-fMRI papers, ranging from 24% to 76%. On average, 53% of checklist items were reported in a given article.

**Conclusions:** Use of the ContES checklist is expected to enhance the methodological reporting quality of future concurrent tES-fMRI studies, and increase methodological transparency and reproducibility.

## Introduction

The advent of functional neuroimaging techniques allows one to investigate the neural correlates of behavior and underlying processes. However, functional neuroimaging techniques cannot by themselves establish causal evidence for brain-behavior relationships. Non-invasive brain stimulation techniques, including low-intensity transcranial electrical stimulation (tES), can be used in combination with functional neuroimaging, such as functional magnetic resonance imaging (fMRI), to directly modulate patterns of neuronal activity and to establish causal evidence for the involvement of particular neural regions or networks in a specific behavior and cognitive process ^1–18^. Over the last 20 years, low intensity tES has been used extensively to study and modulate the neural mechanisms underlying basic physiological and cognitive processes ^19–27^. Initial studies combining tES with fMRI were limited to sequential tES-fMRI recording, which primarily provides an avenue to investigate the neural mechanisms underlying tES offline (after) effects ^28–34^.

Over the last decade, advances in tES technology have made concurrent tES-fMRI (i.e., simultaneous acquisition of fMRI data during tES) in-principle technically feasible, thus enabling monitoring of immediate (online) tES effects. Concurrent tES-fMRI recording poses specific technical challenges ^35^, however, these issues can be minimized when standard protocols are followed ^29,36–39^. As a “perturb-and-measure” approach ^40^, applications are rapidly diversifying such that concurrent tES-fMRI is being used increasingly as a proxy measure for local and global neuro-metabolic activity to address causal mechanistic ^25,41,42^ and predictive ^43,44^ questions about underlying physiology and therapeutic effects. Online integration of tES with fMRI recordings is, however, associated with technical and theoretical challenges, which include the risk of electrode heating due to the radio frequency (RF) pulses of the scanner ^45,46^ and susceptibility-related echo-planar imaging (EPI) artifacts under the electrodes ^36,47^. Furthermore, evidence is increasing for the significant impact of different methodological procedures on online fMRI responses to tES, including the localization of electrodes ^47–49^, MRI-conditional stimulator setup ^29,37,38^, amount/type of contact medium ^35^, and the timing of concurrent tES within the fMRI paradigm ^50,51^. Given the variability in fMRI responses to tES, as well as tolerability/safety/noise concerns and methodological variations, there is an urgent need to clearly and systematically plan, measure, report, and control as many of these methodological factors as possible. In order to ensure a robust interpretation of the data and to increase the potential for future replication, a reporting guideline and checklist is required. Methodological checklists not only improve the transparent reporting of study methodology, and quality of data collection analysis, but also reduce design and reporting biases, factors with clear implications for future interpretation and use of the data. These checklists could also assist peer review and critical appraisal of research methodology ^52–54^.

A limited number of methodological checklists are available in the field of human brain mapping for transcranial magnetic stimulation (TMS) studies ^53^, tDCS studies ^54^, and MRI/fMRI studies ^55,56^. One of the most well-known checklists in the field of human brain mapping is the COBIDAS statement, which was developed to provide an evidence-based minimum set of recommendations to prepare best practices for data analysis, result reporting, algorithms, and data sharing in neuroimaging research in order to promote transparency, reliability, and collaboration ^55^. Given the potential for variability of the neural responses elicited by tES and the growing number of concurrent tES-fMRI studies, guidelines on factors that should be reported and/or controlled in concurrent tES-fMRI studies are essential to ensure that research findings are correctly interpreted and reproducible ^57^. Also, to facilitate meta-analyses, studies should be consistent in both methodology and reporting practice. Hence, we aimed to address these issues by conducting a Delphi study to reach a consensus on the essential items that are mandatory to be reported or recommendations that should be considered when reporting a concurrent tES-fMRI study (ContES Checklist).

## Research Methodology

The Delphi method is a questionnaire-based approach designed to facilitate reaching a consensus, based on the fundamental principles of purposive sampling of experts in the field of interest, panelist anonymity, iterative questionnaire presentation, and feedback of statistical analysis ^58–60^. Like other expert consensus methods, the Delphi method is sensitive to expert sampling and opinion aggregation choices and is reliant on subjective expert judgement inherently, necessitating the use of other complementary empirical evidence ^60,61^. However, rigorously collected and synthesized expert opinion constitutes an important source of information when empirical data is scarce and issues of interest are complex and multifaceted ^62,63^.

This study was designed, implemented, and coordinated within the international network of tES-fMRI (INTF) and a steering committee (SC) that supervised the process of checklist development, data analysis, and determining the initial criteria for item consensus and survey termination. The flowchart of the Delphi method adapted for this study is illustrated in Figure 1. The development of the ContES checklist using the Delphi technique involved the following steps: (1) formation of the SC, (2) selection of the expert panel (EP), (3) checklist development and revision, and (4) data collection and analysis. The protocol of this study is pre-registered in OSF ^64^ and its questionnaires and databases are publicly available in the study’s OSF page (https://osf.io/f9j8z/).

**Figure 1.**
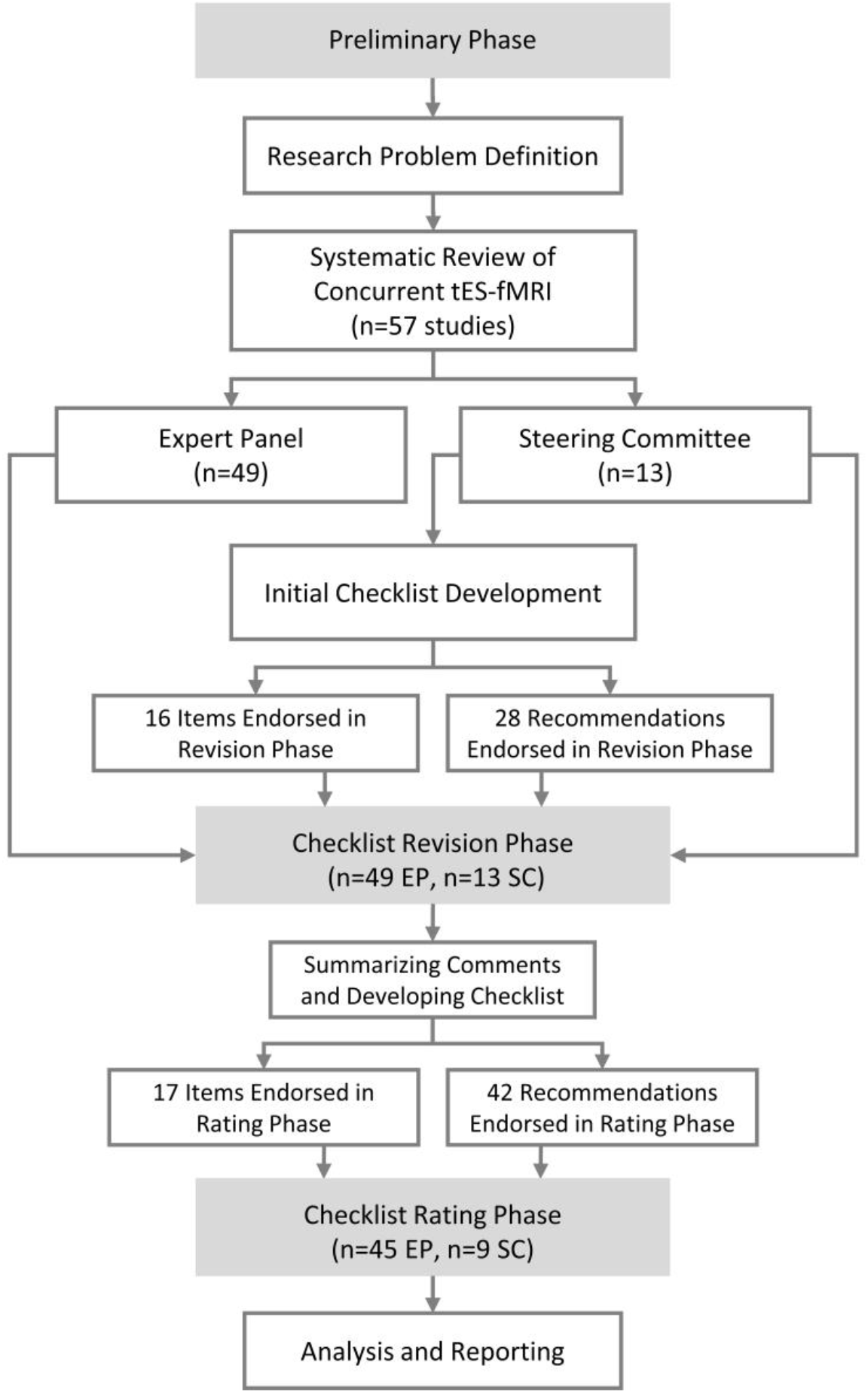
Flowchart diagram of the Delphi process to develop the checklist. The Delphi process started with members of the steering committee defining the research problem. Then the field of concurrent tES-fMRI studies was systematically explored to find eligible people to invite to the steering committee and expert panel. The checklist was then developed by the steering committee and then was sent for revisions to the expert panel. After this phase the checklist was revised by the steering committee and then was sent for the rating phase. At the final stage, the ratings were analyzed. “n” indicates the number of participants in each group.

### Steering Committee (SC)

The role of the SC, comprising Jorge Almeida, Andrea Antal, Marom Bikson, Hamed Ekhtiari, Lucia M. Li, Marcus Meinzer, Michael Nitsche, Duke Shereen, Hartwig Siebner, Charlotte Stagg, Axel Thielscher, Ines Violante, and Adam Woods, was to determine the aim of the research, produce items and select additional experts for the Delphi process. Peyman Ghobadi-Azbari served as the Delphi facilitator to implement the preregistered methods within and between the SC and expert panel (EP). The SC grew out of the INTF collaborative group after a series of webinars (28 March 2019, 27 June 2019, and 26 September 2019; recorded videos of the webinars are available on the INTF YouTube channel https://youtube.com/channel/UCKcEYDmyqTipDW7OzuoVSlg), in which considerable heterogeneities of technical/methodological aspects in studies combining tES with fMRI were discussed along with strategies to help to bridge respective knowledge gaps, and reduce heterogeneity.

### Expert Panel (EP)

The project involved the recruitment of a group of experts based on a systematic review of 57 concurrent tES-fMRI studies (published before January 1^st^, 2020). We reviewed the concurrent tES-fMRI literature in the PubMed research database from inception up to January 1, 2020 to select evidence based concurrent tES-fMRI studies and experts who conducted those studies. The Preferred Reporting Items for Systematic Reviews and Meta-Analyses (PRISMA) ^65^ flow diagram for the systematic review is provided in Extended Data Fig. 1. The search included the terms (*tDCS* OR *transcranial direct current stimulation* OR *tACS* OR *transcranial alternating current stimulation*) AND (*functional magnetic resonance imaging* OR *fMRI* OR *functional MRI* OR *fcMRI* OR *functional connectivity MRI* OR *rsfMRI* OR *resting-state fMRI*). Fifty-seven articles were selected based on the PRISMA. The inclusion criteria used to invite the experts included being the first, last or corresponding author in at least one of 57 published studies in the field. In addition, the members of the SC were asked to nominate additional experts in the field of concurrent tES-fMRI to join the EP. All SC members agreed on the list of experts before the invitation process was started. Potential candidates for the EP based on the above-mentioned inclusion criteria (n=54) were invited to participate in the Delphi study using the contact information provided in each publication (the e-mail address of the respective contributor). Furthermore, the committee invited 21 additional experts to join the EP. The final list of EP invitees included 75 potential candidates with expertise across a range of backgrounds (i.e., medicine, neuroscience, biomedical and electrical engineering) and geographical areas (USA, UK, Germany, Denmark, Iran, and Canada). Over 65% of the invitees (49 experts) accepted to join the EP.

### Checklist Development Phase

The checklist aimed to facilitate an in-depth consensus among the tES-fMRI experts regarding the technical/methodological aspects necessary to be followed and reported to safely and successfully perform acquisition of fMRI during tES delivery and to enable critical appraisal and systematic reporting of concurrent tES-fMRI studies. The initial draft of the checklist was developed based on currently available evidence in the field. The concurrent tES-fMRI studies were operationally defined as “studies that apply tES in the bore of the magnet while acquiring fMRI data during stimulation”. Studies using tES-fMRI in offline or sequential approaches (i.e., imaging only before and after stimulation) to evaluate the short- and long-term after-effects of brain stimulation were not included. As the first step of the Delphi process, an initial email circulation started within the SC by asking each SC member to suggest a list of the specific technical/methodological aspects of the interaction between fMRI and tES that they considered very likely to influence a concurrent tES-fMRI study and its report. Repeated responses were merged and the remaining items were thematically categorized into: technological factors, safety and noise tests, and methodological factors. The SC also suggested additional recommendations for each main item that should be considered in order to increase the quality of reporting. Following agreement on the checklist format by the SC, the initial draft of the checklist was tested by rating 5 sample concurrent tES-fMRI articles, with Yes/No ratings on whether the item was reported in the article or not, to ensure the checklist’s objectivity and clarity. Following the pilot test, the SC reworded and/or combined items that were deemed unclear for inclusion in the revision phase. The results of each phase were summarized and displayed on the study’s OSF page (https://osf.io/f9j8z/).

### Data Collection and Analysis

#### Checklist Revision Phase

The consensus-based checklist was distributed among the EP and SC members. For the revision phase, contributors were sent the initial checklist email. Two consecutive follow-up reminders were emailed if a response was not received after 7 and 14 days following the initial email circulation. Contributors who completed the revision phase before the deadline were recruited in the subsequent rating phase. The revision phase included a section on self-reporting the demographics gleaned from the EP and SC members and questions about their previous experiences as concurrent tES-fMRI researchers. A second section requested contributors to comment on any ambiguity or wording of the existing checklist. The revision phase included a definition of the purpose of the consensus study and an operational definition of a prescriptive standard protocol for concurrent tES-fMRI trials, the presentation of the initial checklist, followed by the opportunity to modify and remove items/recommendations, revise the current language of the checklist, merge selected items/recommendations, and propose new items/recommendations for each subsection. Any item that was judged by the SC as an original idea was included as a new item/recommendation in the rating phase. Data obtained from the revision phase informed the SC in developing the finalized checklist.

#### Checklist Rating Phase

In the rating phase, the EP and SC members were sent a feedback document, which summarized the results of the checklist modifications. It included the clarification and correction of terminology, as well as a summary of comments. The participants were asked to rate each item in terms of importance in the methodology of concurrent tES fMRI studies, from 1 to 5. The exact question was: “To facilitate visibility, replication and data sharing, how important is it to report this item?”. Also, for each additional recommendation, we asked: “Do you support the inclusion of this additional note as a recommendation to be considered in concurrent tES fMRI studies?”.

To avoid a non-neutral center rating and encourage deliberation, ratings were termed “not important”, “slightly important”, “moderately important”, “highly important” and “extremely important”. The participants were also allowed not to rate an item if they chose not to do so. The inclusion of each additional recommendation for each item could be rated “Yes” or “No”.

#### Data Analysis

In the rating phase, the average rating and the number of responses were calculated. For the main items, the tally of scores of “extremely important”, “highly important” and “moderately important” represented “essential”, whereas the tally of the scores of “slightly important” and “not important” represented “non-essential”. We defined consensus as ≥70% of respondent scorings of an item as essential, with a second, preferred level of consensus at ≥80% agreement. Also, for additional recommendations, all respondents rated the 42 recommendations with the scores of “Yes” or “No”, as previously described. The recommendation items receiving a response of “Yes” from at least 50% of EP and SC members were defined as achieving consensus.

#### Assessing the State of Reproducibility and Transparency in concurrent tES-fMRI studies with the ContES Checklist

To retrospectively assess the state of reproducibility and transparency in reporting via adherence to the ContES Checklist in published concurrent tES-fMRI studies, we evaluated 57 studies using the ContES checklist. Three independent raters (HT, NM, HMA) rated adherence to the reporting checklist within these articles using the 17-items checklist. An inter-rater reliability analysis using the Fleiss’ Kappa statistic was performed to assess the consistency of the raters’ evaluations of concurrent tES-fMRI research in the context of the ContES checklist ^66^. If Fleiss’s Kappa is greater than 0.8, the accuracy of the inter-rater reliability indicates “Almost Perfect Agreement” ^67^. The relationship of reporting score with publication year, journal word limit, article word count, and journal impact factor were also analyzed to assess whether articles with a better reporting status appear in journals with higher impact factors, whether the reporting status has improved across the recent years, and whether word count limitations have an impact on reporting status. None of these relationships were significant. Also, the number of example articles reporting each item is presented in Supplementary Table 1. To support the potential utility of the checklist, this table also provides a list of papers that demonstrates how each checklist item might affect the results of a concurrent tES-fMRI study as well as their importance for interpretability and generalizability. A summary of these 57 concurrent tES-fMRI studies is provided in Supplementary Table 2.

### Ethics

No ethics board approval was required for this expert panel activity. This consensus-seeking activity neither involved novel experimental work nor novel analyses of existing experimental data, but relied entirely on mutual exchange of expertise and opinions within the panel taking into account all existing peer-reviewed scientific studies on concurrent tES-fMRI. Potential contributors were informed that by responding to the invitation letter, they were deemed to have consented to take part in the Delphi study and that their de-identified responses are included in all analyses. All named contributors also provided consent to be acknowledged in this paper.

## Results

### Characteristics of the Steering Committee and Expert Panel and Response Rates

The characteristics of the SC and EP are presented in Supplementary Table 3. The SC and EP had a mean (SD) of 8.67 (5.4) and 5.54 (2.7) years of experience in tES-fMRI research, respectively. They represented a range of professions and academic disciplines including neuroscientists (49% EP, 85% SC), cognitive scientists (16% EP, 8% SC), psychiatrists (10% EP, 0% SC), and psychologists (10% EP, 0% SC). Their professional settings were primarily universities (59% EP, 69% SC), hospitals (18% EP, 8% SC), university hospitals (0% EP, 8% SC), independent research institutes (6% EP, 15% SC), and businesses/industries (6% EP, 0% SC), and the most commonly held academic degrees were PhD (76% EP, 69% SC), MD-PhD (6% EP, 15% SC), and MD (4% EP, 15% SC). Forty-nine EP members, along with 13 SC members, (completed the revision phase of the Delphi questionnaire and 45 EP members and 9 SC members completed the rating phase. Retention was very high, with 54 (87.1%) revision phase contributors also completing the rating phase.

### Results of the Delphi Process

#### Checklist Development Phase

Four members of the SC (ADS, IRV, JA, HE) produced an initial list of items for the overall structure of the checklist based on suggestions derived from the concurrent tES-fMRI studies literature. After the discussions within the SC, the checklist was expanded from 14 items to 16 items. Thus, for the revision phase, 9 items in the *Technological Factors* category, 4 items in the *Safety and Noise Tests* category, and 3 items in the *Methodological Factors* category were provided within the checklist. Furthermore, an “Additional Recommendations” column was added to the ContES checklist by the SC with 28 additional recommendations for experimental parameters and practices. These additional recommendations provide guidance to the requirements for adequate, and appropriately documented simultaneous conduction of fMRI and tES.

#### Checklist Revision Phase

In the revision phase, one item was added to the ContES checklist (tES-fMRI Setting Test - Subjective Intolerance Reporting). The additional recommendations were expanded by the contributors from 28 items to 42 items. The final checklist includes 9 items and 19 recommendations in the *Technological Factors* category, 5 items and 12 recommendations in the *Safety and Noise Tests* category, 3 items and 9 recommendations in the *Methodological Factors* category as well as 2 general recommendations. Different versions of the checklist in its development process are provided by the study’s OSF page (https://osf.io/f9j8z/).

#### Checklist Rating Phase

The collected responses of the rating phase are shown in Figures 2 and 3, and also in Tables 1, 2 and 3. Respondents had a high rate of agreement about most of the checklist items. However, three items (marked with † in Figure 2): *Amount of Contact Medium (Paste/Gel/Electrolyte), Electrode Placement Visualization*, and *Wire Routing Pattern* did not reach the 80% consensus threshold (rated as either moderately, highly, or extremely important by more than 80% of the respondents). Of these, one item, *Amount of Contact Medium (Paste/Gel/Electrolyte)*, did not reach the ≥70% consensus (marked with ‡ in Figure 2 and Supplementary Table 4). However, the draft ContES checklist met the consensus level for all 17 items with a 65% threshold. The rating phase included also scoring of each of the additional recommended items by the scoring choices of Yes and No. The results showed that 38 (90%) of the recommendations reached the 50% threshold (rated as Yes by more than 50% of the contributors), but the following 4 recommendations did not (10%) (Figure 3): *Control of Amount of Contact Medium, Attenuation Characteristic of RF Filter, Restrictions/Regulations for RF Filtering Method*, and *Restrictions/Regulations for Wire Routing Pattern*.

**Figure 2.**
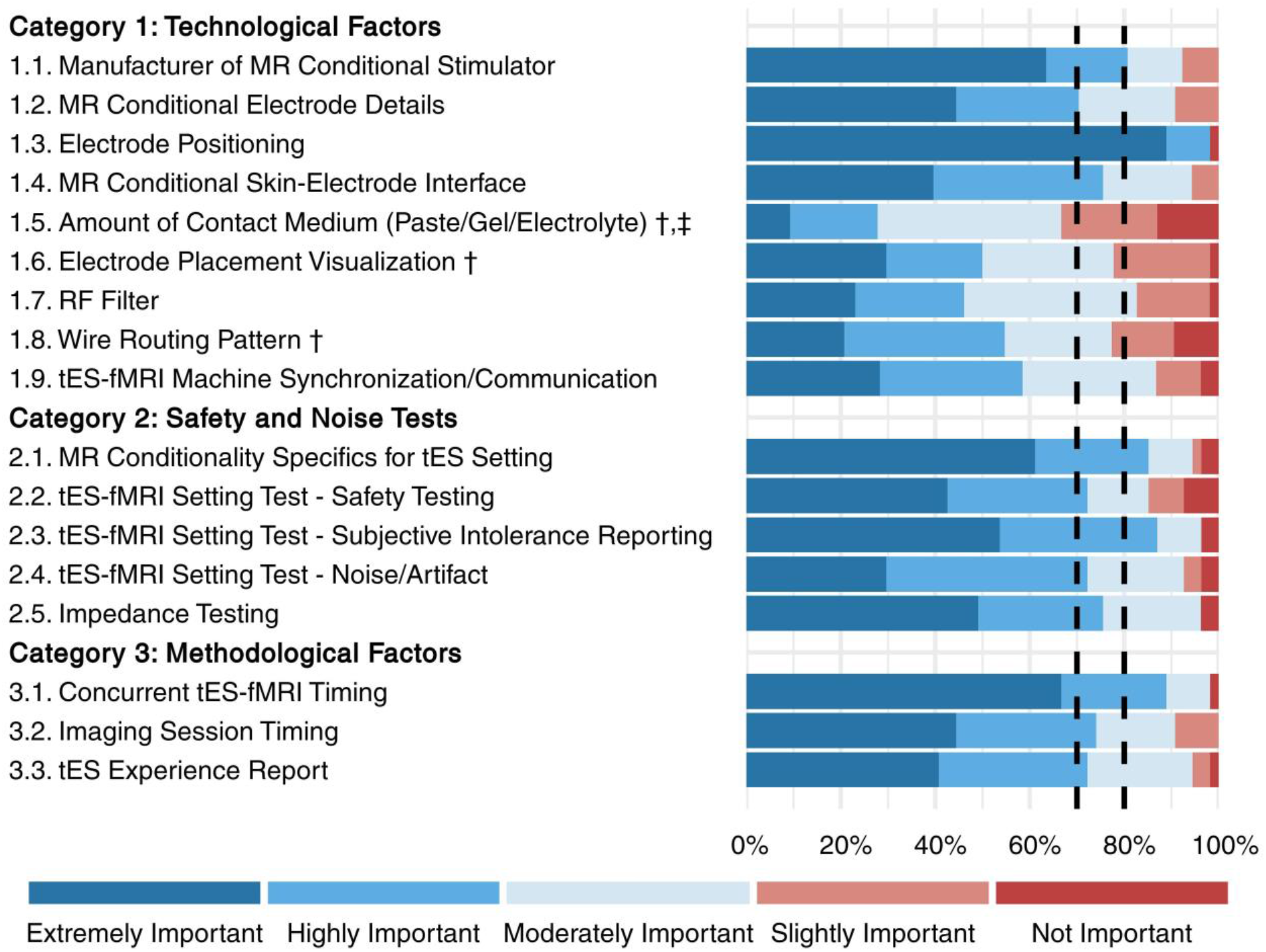
Collected responses from contributors regarding the importance of the main items (rating phase). This figure depicts the rating of the checklist items by 54 respondents in the rating phase. Each item was rated from 1-5 (not important-extremely important). 14 items reached the 80% threshold (rated as either moderately, highly, or extremely important by more than 80% of the respondents). The items that did not reach this threshold are marked with “†”). 16 items reached the 70% threshold (rated as either moderately, highly, or extremely important by more than 70% of the respondents). The one item, which did not reach this threshold is marked with “‡”. Full text of the items is provided in Tables 1-3.

**Figure 3.**
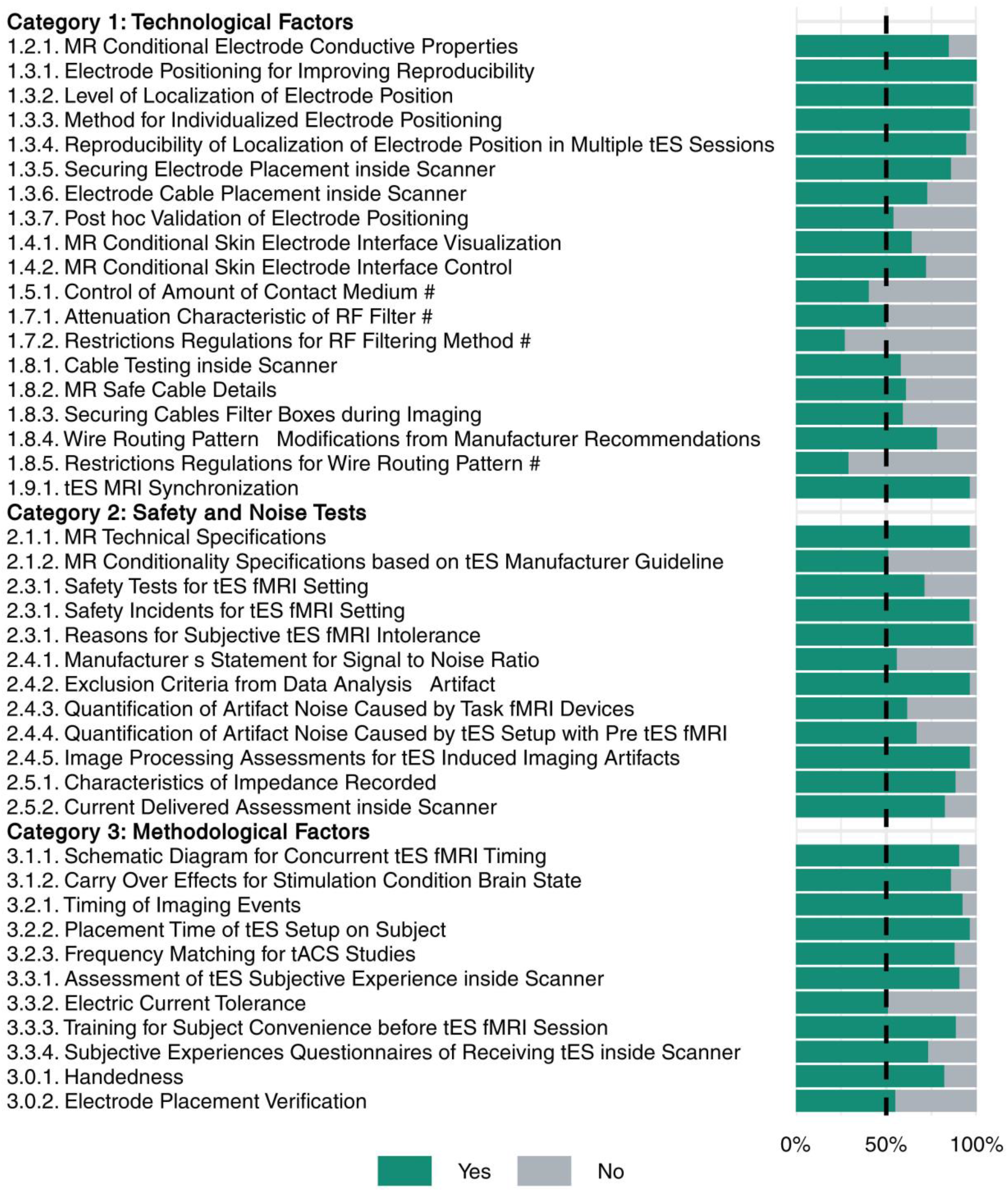
Collected responses of the contributors regarding the importance of recommendations (rating phase). Each additional recommendation was rated either “Yes” or “No” with respect to the question of whether it should be included as a recommendation. The recommendations rated with Yes by fewer than 50% of the respondents are marked with “#”. Recommendations are represented by their summary in the figure. Full text of the recommendations is provided in Tables 1-3.

**Table 1.**
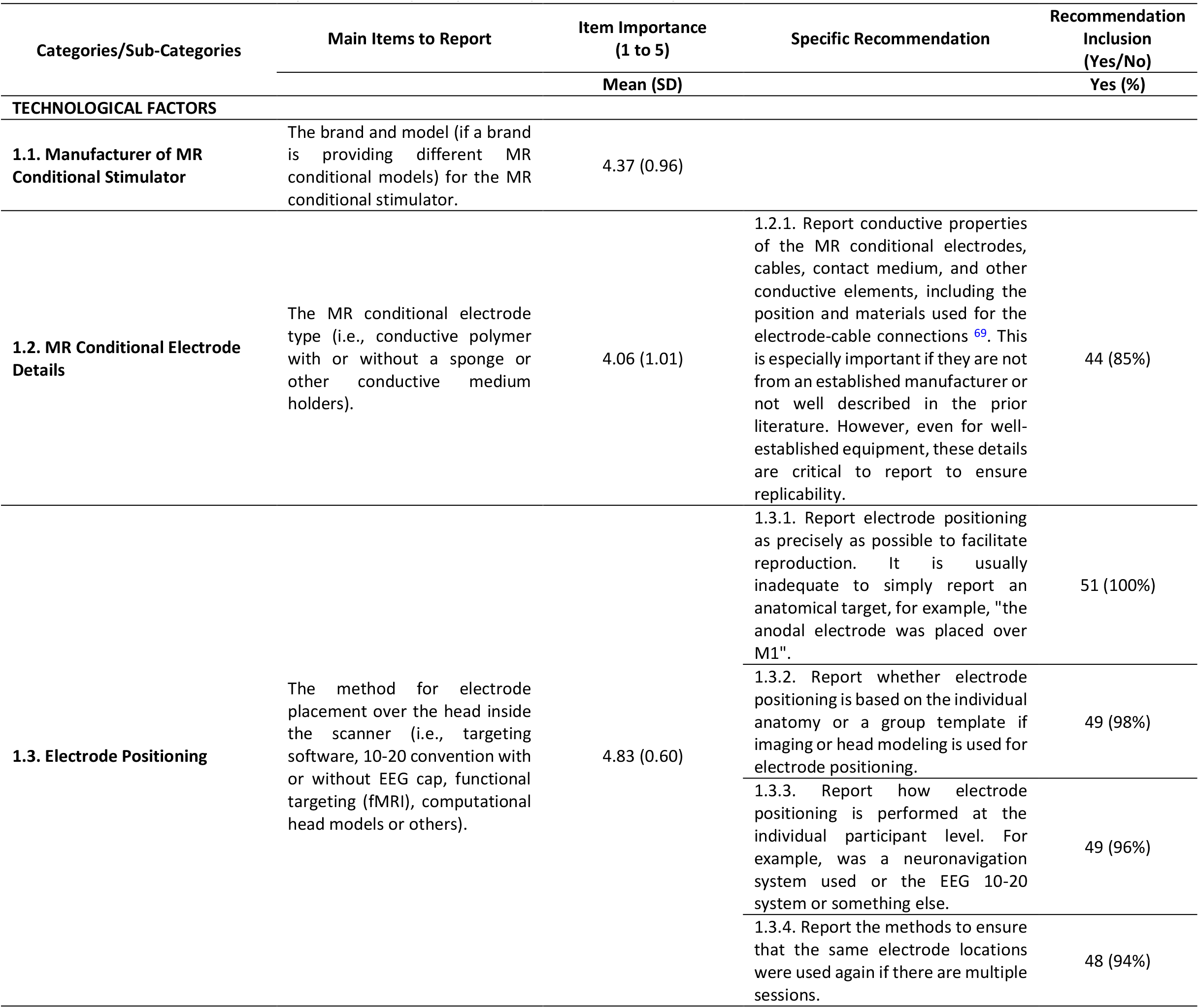
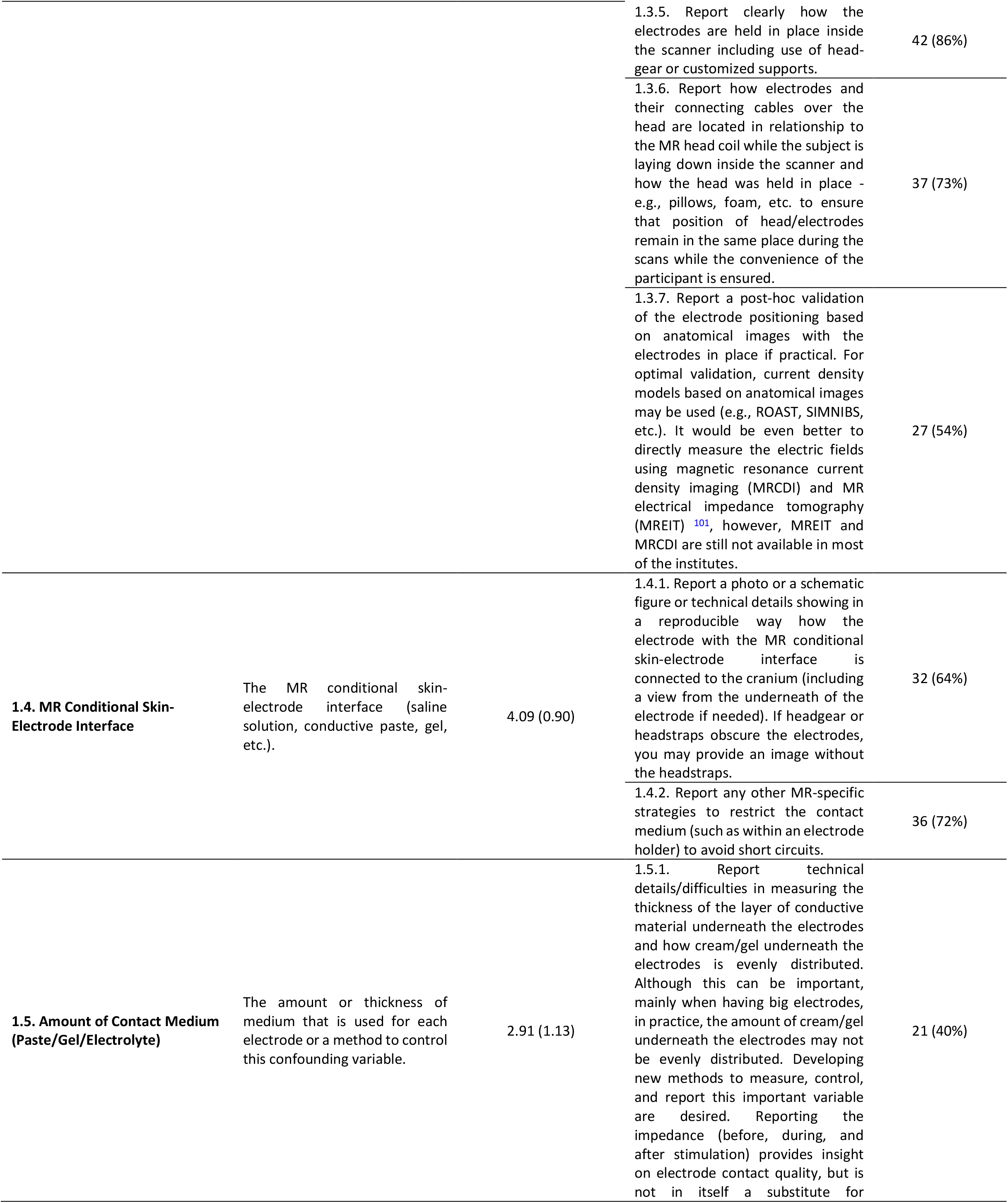
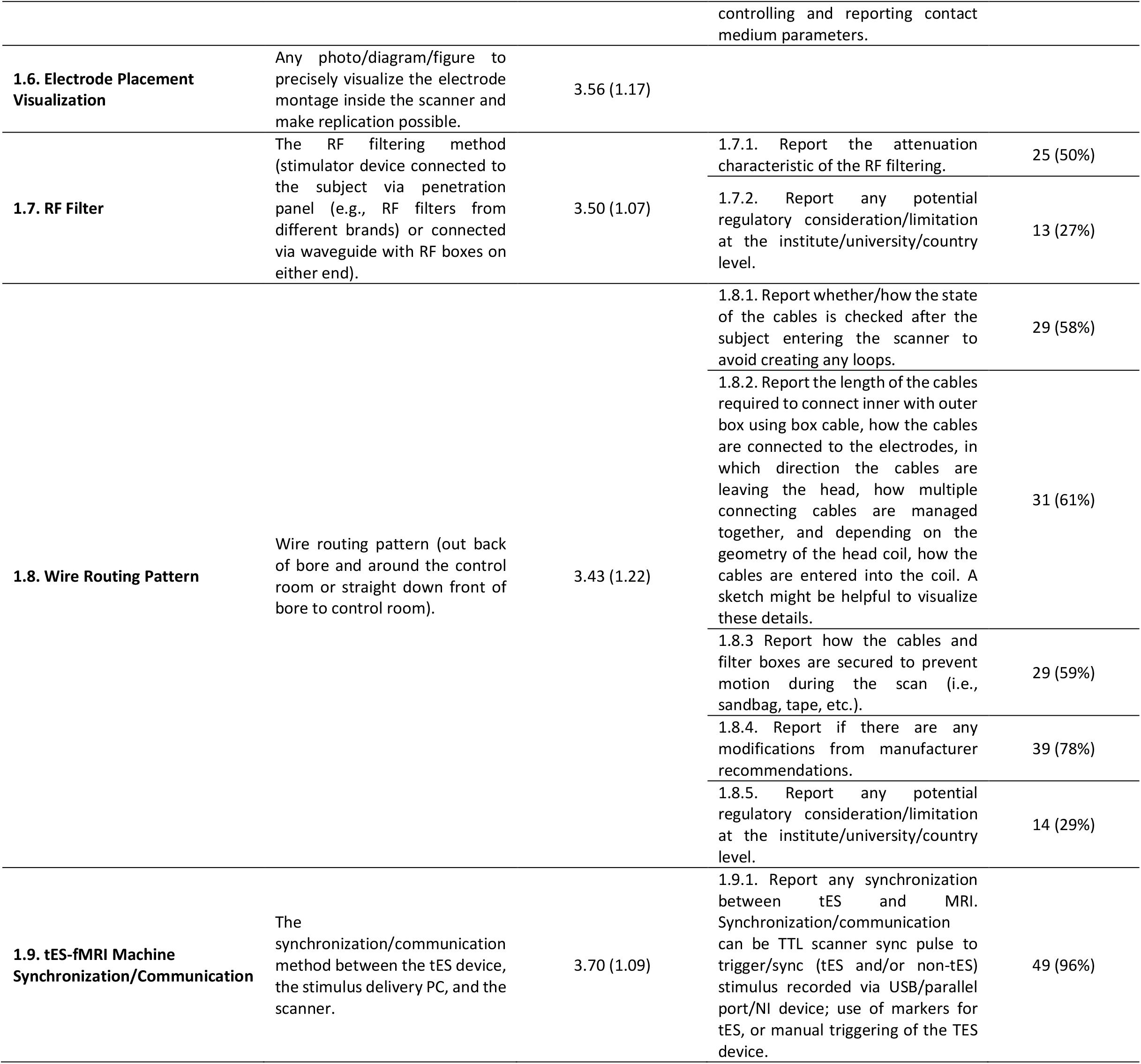
Concurrent tES-fMRI (ContES 2021) Checklist: main items and recommendations of the TECHNOLOGICAL FACTORS category to report in concurrent tES-fMRI research. Ratings for items (scores 1-5) are reported as mean (standard deviation) and ratings for recommendations (Yes/No) are reported as frequency of Yes (percent of Yes reports).

**Table 2.**
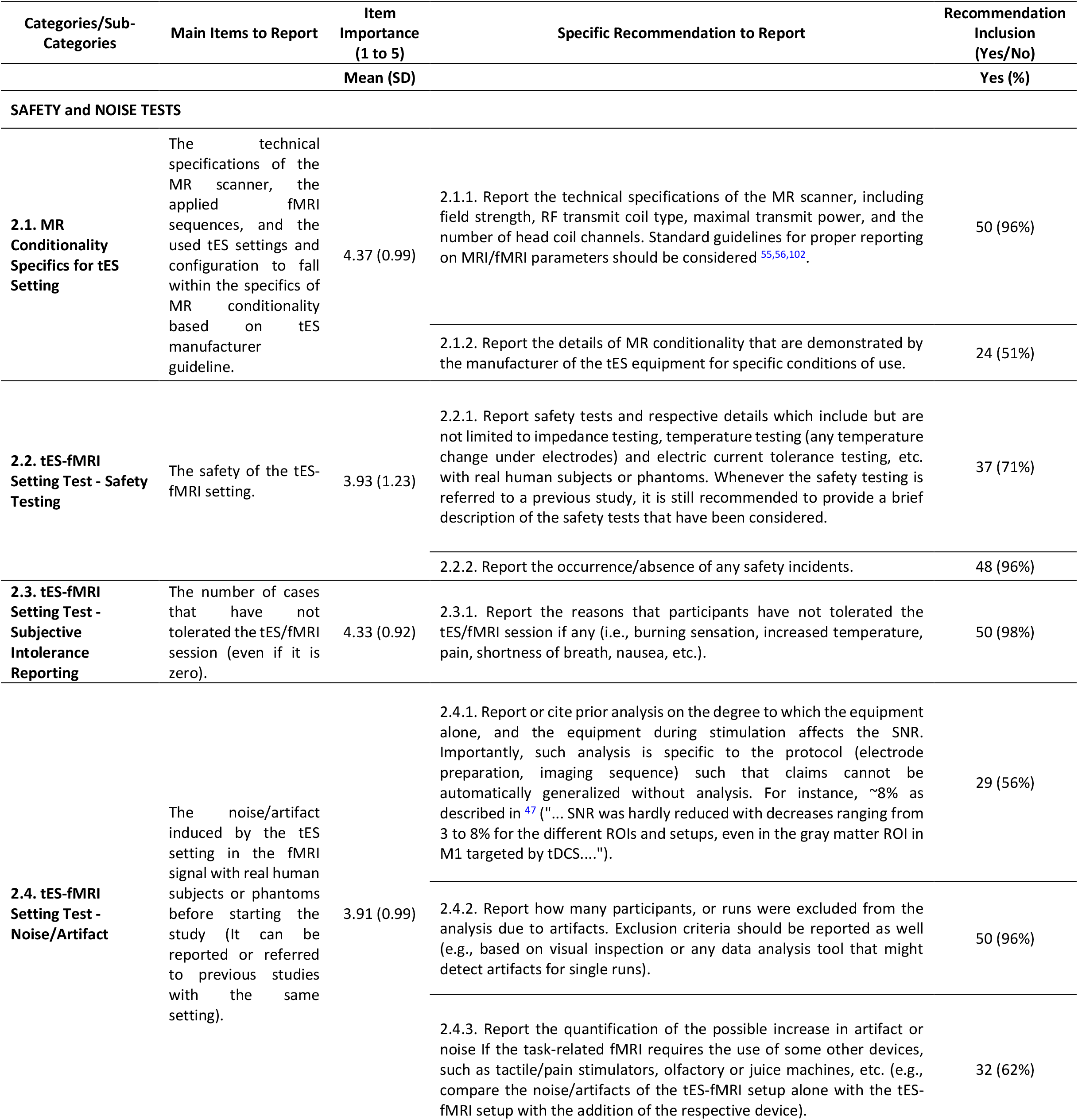
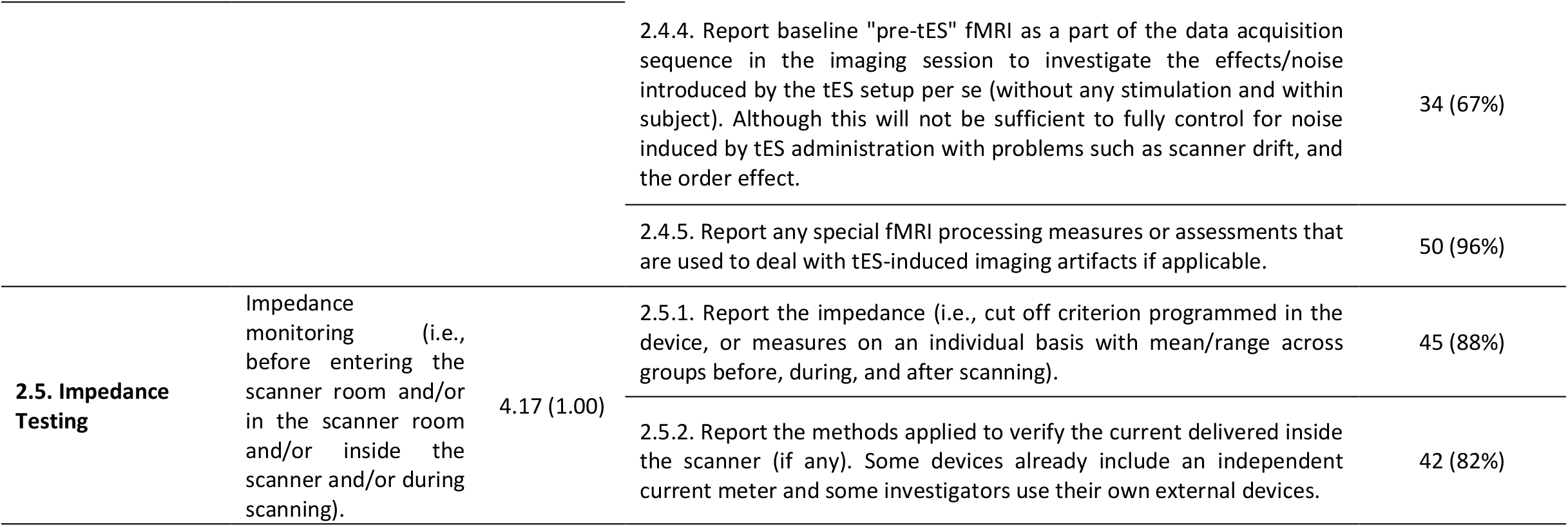
Concurrent tES-fMRI (ContES 2021) Checklist: main items and recommendations of the SAFETY and NOISE TESTS category to report in concurrent tES-fMRI research. Ratings for items (scores 1-5) are reported as mean (standard deviation) and ratings for recommendations (Yes/No) are reported as frequency of Yes (percent of Yes reports).

**Table 3.**
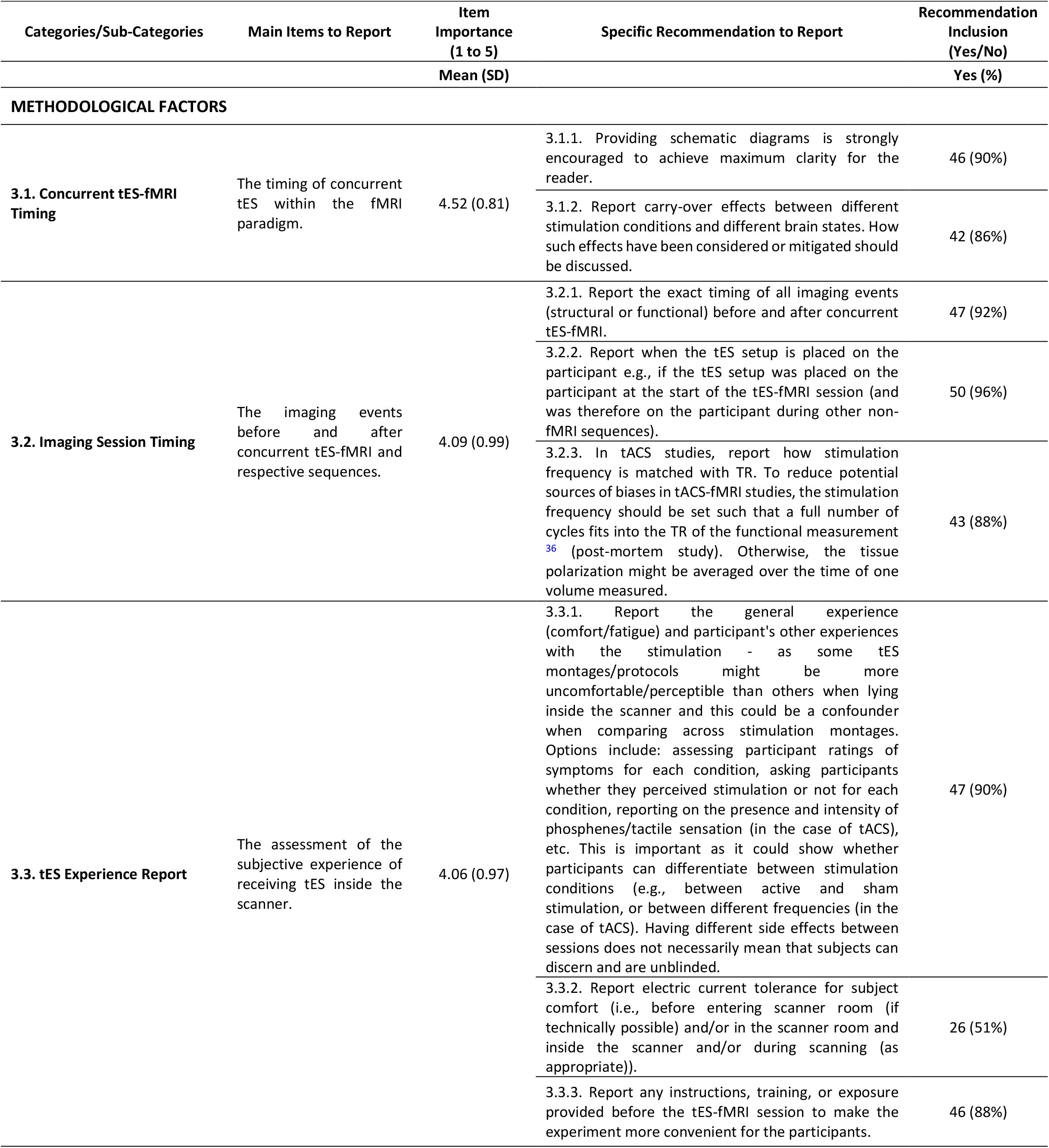
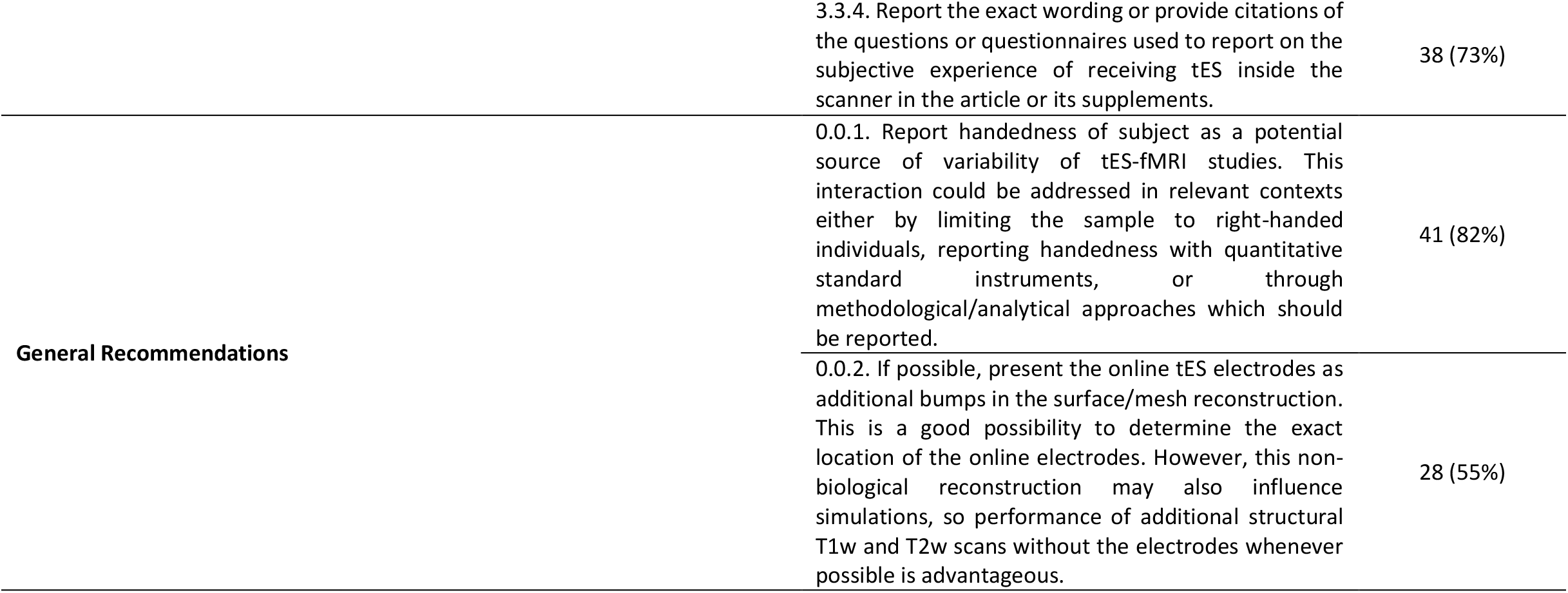
Concurrent tES-fMRI (ContES 2021) Checklist: main items and recommendations of the METHODOLOGICAL FACTORS category to report in concurrent tES-fMRI research. Ratings for items (scores 1-5) are reported as mean (standard deviation) and ratings for recommendations (Yes/No) are reported as frequency of Yes (percent of Yes reports).

The ratings of the items and recommendations of the ContES checklist are outlined in Tables 1-3. The full version of the ContES checklist that includes 17 essential items and 42 additional recommendations and a short version that includes essential items only are provided in Supplementary Tables 4 to 7 to be used by authors and reviewers. The reporting items that did not meet the 70% and 80% thresholds and additional recommendations that did not meet the 50% thresholds are marked in the final checklist. Based on this information, researchers can decide to choose more stringent or more liberal thresholds when using the checklist.

#### The State of Reproducibility and Transparency in concurrent tES-fMRI studies with the ContES Checklist

Three independent raters evaluated the adherence of the concurrent tES-fMRI articles to the finalized reporting checklist items. The consistency of the raters’ responses resulted in a Fleiss’ Kappa of 0.85, indicating that the consistency is *Almost Perfect Agreement* ^67^.

Inclusion of information about the main items of the ContES checklist varied widely, ranging from fully reported (100%; Manufacturer of MR Conditional Stimulator, Concurrent tES-fMRI Timing, Imaging Session Timing) to rarely reported (5.3%; MR Conditionality Specifics for tES Setting, Amount of Contact Medium). The pattern of adherence to the checklist items varied relevantly between articles, ranging from 24% to 76%, averaging 53% of checklist items reported in a given article (Figure 4).

**Figure 4.**
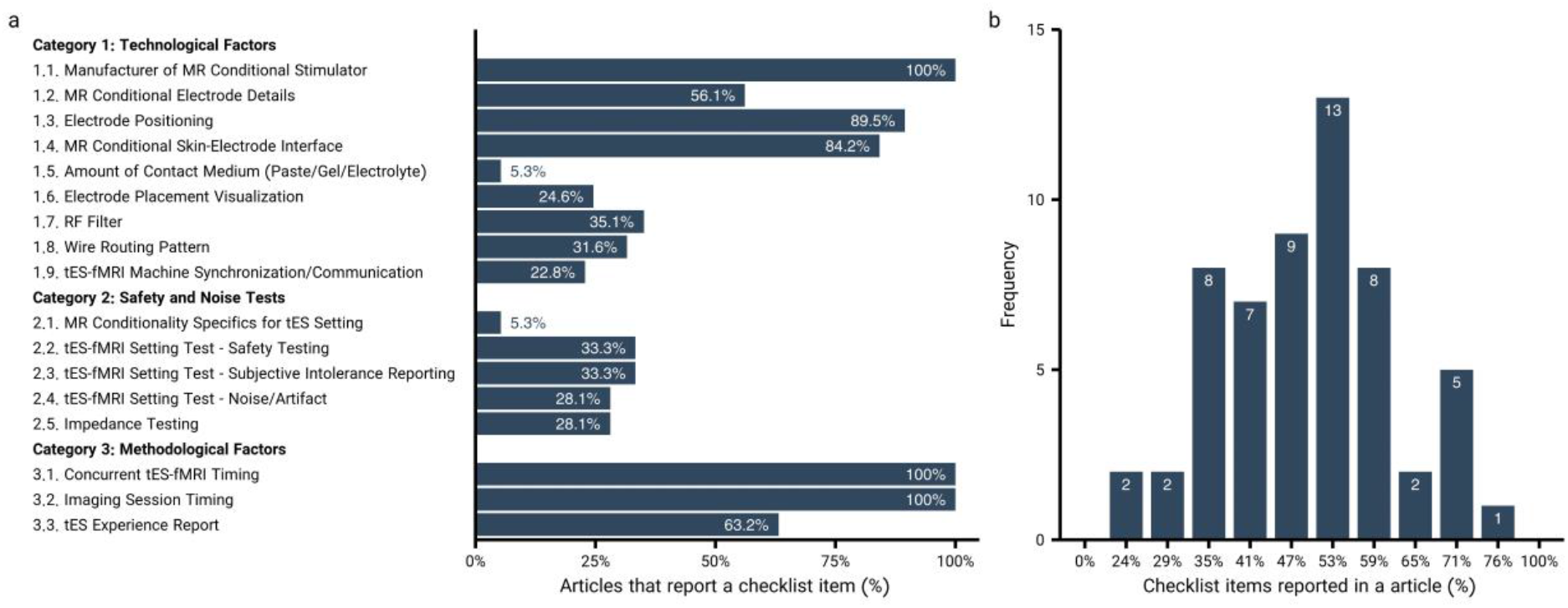
State of reproducibility/transparency in concurrent tES-fMRI research in the context of the ContES checklist. Assessments by 3 independent raters are based on 57 tES-fMRI papers, from the first published study up to January 1, 2020. **a**, Percentage of the articles that adhered to each checklist item. **b**, The checklist items adhered to by the 57 articles.

All studies (100%) reported the Manufacturer of MR Conditional Stimulator (Item 1), and the Electrode Positioning (Item 3) was described clearly in 89% of articles, but details of the MR Conditional Electrode (Item 2) were included in only 56% of the reviewed articles. A relatively high number of papers (84%) reported the MR Conditional Skin-Electrode Interface (Item 4), but the Amount of Contact Medium (Item 5) was mentioned less frequently (5%). The Electrode Placement Visualization (Item 6) was shown in only 25% of the articles, and the RF Filter (Item 7) was included in 35% of the articles. The Wire Routing Pattern (Item 8) was described clearly in only 32% of articles, and the tES-fMRI Machine Synchronization/Communication (Item 9) was rarely described (23%).

Only 5% of the articles reported information regarding MR Conditionality Specifics for tES Setting (Item 10). Few articles described details of the tES-fMRI setting test, ranging between 28 and 33% in Items 11-13 (33% Safety Testing, 33% Subjective Intolerance Reporting, 28% Noise/Artifact). Impedance Testing (Item14) information was included in only 28% of articles. Concurrent tES-fMRI Timing (Item 15) and the Imaging Session Timing (Item 16) were reported in all 57 articles, however tES Experience was reported less frequently (Item 17; 63%).

The highest reporting score was 76%, and 6 articles had a score of higher than 70%. One article reported more than 75% of the checklist items ^68^. The lowest reporting score was 24%; 28 studies failed to meet a reporting threshold of 50%. The correlations of study reporting status with journal word limit, article word count, and journal impact factor were not significant and relevant graphs are presented in Extended Data Fig. 2.

## Discussion

The goal of this study was to develop a consensus-based checklist of methodological details to facilitate the evaluation of concurrent tES-fMRI studies in terms of methodological transparency and reproducibility (ContES Checklist). We successfully developed the ContES checklist to guide authors in reporting the minimum information necessary to ensure reproducibility by using the 17 essential items. The 42 additional recommendations should be considered to further enhance the quality of future research in this field. This checklist can be used by editors and reviewers for critical appraisal of future studies. The checklist will also be helpful for researchers who are in the process of setting-up a concurrent tES-fMRI study. Indeed, our systematic literature review and appraisal of 57 published concurrent tES-fMRI studies revealed a general lack of sufficient information to fully reproduce critical methodological details of these studies. Overall, this checklist offers a methodological framework for understanding and replicating previous studies, and provides journal reviewers and editors with an efficient tool to gauge and promote concurrent tES-fMRI reproducibility. Figure 5 summarizes the items which are deemed important to be considered when conducting and reporting a concurrent tES-fMRI study.

**Figure 5.**
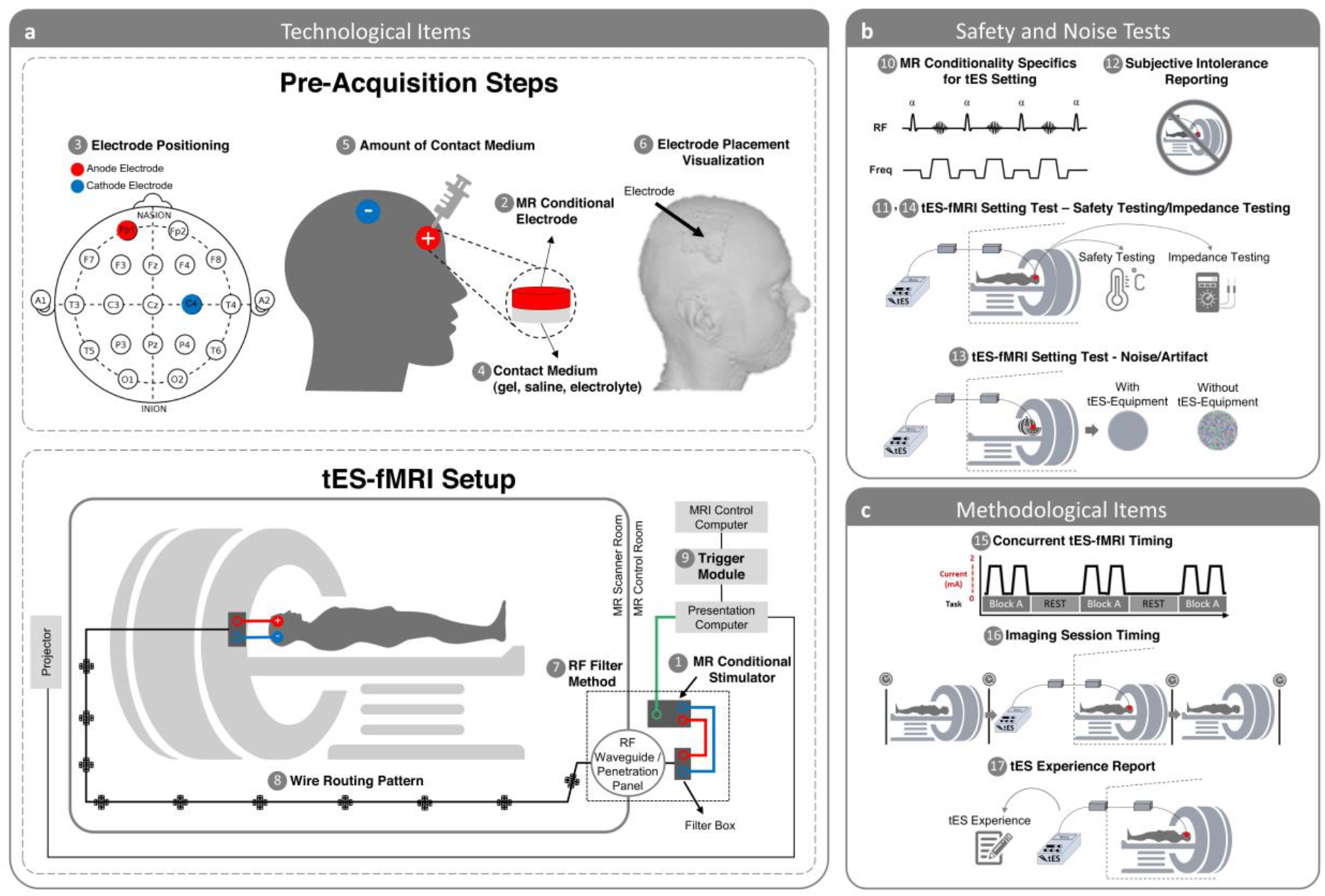
Scheme of the concurrent tES-fMRI approach in the context of the ContES checklist. **(a) Summary of technological considerations.** MR conditional stimulator (1, item 1.1) is connected to the head through RF waveguide or RF penetration panel (7, item 1.7). Box cable should be aligned with the wall of the scanner room and run parallel to the bore axis (8, item 1.8). MR conditional stimulator is connected to the outer filter box or RF band-stop filter adapter as well as to the presentation computer trigger output cable. Synchronization module (9, item 1.9) should be connected to the presentation computer as well as to the MRI control computer. Electrode positioning (3, item 1.3) is used to accurately stimulate cortical target regions and exert neuromodulatory effects. A method allowing quantification of contact medium (e.g., syringes) should be used to achieve a consistent and appropriate amount of contact medium (5, item 1.5). MR conditional skin-electrode (e.g., saline solution, conductive paste, gel) (2, item 1.2) is used to facilitate delivery of current to the scalp (4, item 1.4). Electrode placement visualization can be used to reproducibly center each electrode on the head so that intrascanner stimulation allows verification of correct positioning of the electrodes on the head (6, item 1.6). **(b) Summary of safety considerations**. MR Conditionality Specifics for tES Setting include the technical specifications of the MR scanner, the applied fMRI sequences, and the used tES settings and configuration to fall within the specifics of MR conditionality based on tES manufacturer guideline (10, item 2.1). The Safety of the tES-fMRI Setting includes electrode temperature testing, electric current tolerance testing, etc. with real human subjects or phantoms (11, item 2.2). tES-fMRI Setting Test – Subjective Intolerance Reporting shows the number of cases that have not tolerated the tES-fMRI session (12, item 2.3). tES-fMRI Setting Test - Noise/Artifact shows the noise/artifact induced by the tES setting in the fMRI signal with real human subjects or phantoms before starting the study (13, item 2.4). The impedance is monitored before entering the scanner room and/or in the scanner room and/or inside the scanner and/or during scanning (14, item 2.5). **(c) Summary of methodological considerations**. Concurrent tES-fMRI Timing shows the timing of concurrent tES within the fMRI paradigm (15, item 3.1). Imaging Session Timing shows the imaging events before and after concurrent tES-fMRI and respective sequences (16, item 3.2). tES Experience Report includes the assessment of the subjective experience of receiving tES inside the scanner (17, item 3.3).

### Technological Factors

The technical features of the stimulator and accessories, including set-up on the subject’s head and configuration inside the scanner, underpin rigor and reproducibility – which in turn informs how these elements should be reported (Table 1). Manufacturer make and model should be reported but the degree to which this satisfies items on the checklist varies. For example, while the material composition of an electrode may not always be explicitly specified, indicating a unique electrode part number would allow reproduction and referencing to other documentation. For other items, the amount of detail given beyond the part number can vary depending on the specific approach. For example, the thickness of paste or amount of other electrolytes is determined by the operator (set-up) for large pad electrodes but controlled by the electrode holder for HD electrodes. The item number does however not explain set-up details such as cable arrangements or ad-hoc steps to support electrode positioning. These aspects are important to state. The degree to which prior papers can be referenced for these methodological details (e.g., “we applied tES-fMRI used methods as reported in these other references”) should be qualified. Our analysis suggests that to date only a limited number of papers documented these details in sufficient detail. To the extent these technical factors underpin reproducibility, expanding on them in any given publication supports rigor.

Electrodes used for stimulation inside the MR environment need to be at minimum MR conditional. Manufacturer and model details, electrode size and shape, as well as materials and conductive properties of electrodes (conductive polymer, Ag/AgCl, etc.), connectors (often residually ferromagnetic), cables, and other conductive materials (e.g., a specific brand of electrode paste and NaCl concentration) need to be provided. The relevant item in the checklist was considered highly important (item 1.2, average rating score: 4.06) and the inclusion of the additional recommendation was recommended by 85% of the contributors (recommendation 1.2.1). Also, the position of the connector on the electrode should be reported, as it can significantly influence the homogeneity of current distribution within the electrode ^69^. As revealed by concurrent tDCS-MEG experiments ^70^, some conductive polymer (rubber) electrodes are magnetized, possibly during the production process, while others from the same brand are not. It remains to be determined whether this property is related to MR imaging artifacts.

Electrode positions and size are crucial parameters which determine the distribution of current flow in the brain tissue. It is therefore recommended unanimously by the experts to report this information (item 1.3, average rating score: 4.83) as precisely as possible. It should be distinguished between how the intended montage is determined and how this is practically implemented. The former may be based on the literature, on theoretical considerations, or dedicated E-field modeling in generic or personalized head models, whereas the latter may involve TMS hotspot-search (for M1), 10-20 EEG system head measurements, or MR-based neuronavigation. The reported details should include the method of electrode positioning (e.g., with or without EEG cap), the position of the electrode center, and if applicable, its orientation in case of non-circularly shaped electrodes. Instead of “the electrode was positioned on the left M1” one would preferably state, for example, that “the electrode was centered on the FDI motor hotspot as determined by TMS, with the longer sides of the 7 × 5 cm^2^ rectangular electrode pointing into anteromedial and posterolateral direction, respectively, and the connector inserted at the center of the electrode pointing towards one shorter side into anteromedial direction”. If MRI-based head modeling was used, it should be stated whether electrode position had been determined based on individual anatomy or a group template, and how electrode positioning was performed, e.g., using a neuronavigation system or EEG 10-20 coordinates. For MR-based neuronavigation, MNI coordinates may be reported for electrode centers and/or corners. In the case of multi-session experiments, measures taken to ensure consistency of electrode placement across sessions have to be described, such as co-registration of stimulation electrodes with the individual MRI using neuronavigation or the use of EEG caps and/or the 10-20 system ^71^. The accuracy of the stimulation montage can only be judged if this information is provided and the detailed information further allows post-hoc current modeling and replication studies.

It is also recommended to report the position of electrodes and, in particular, how the cables are directed (intertwined or separated) relative to the MR head coil, as well as information on how electrodes were affixed to the head in the MR, and how the head was stabilized to prevent movement of electrodes relative to either head or MR coil during recordings to prevent discomfort, impedance issues, and imaging artifacts, respectively. A post-hoc validation of electrode positions can also be achieved by the acquisition of anatomical images with the stimulation electrodes in place, even though such images might not be easy to use for E-field modelling itself (due to the challenges of segmentation between the electrodes and skin), for which anatomical images without electrodes (and related artifacts) are preferred.

For the sake of reproducibility, it is also important to provide a proper visualization of electrode position (item 1.6, average rating score: 3.56), which may be a photo, a sufficiently detailed schematic figure, or, preferably, the precisely modeled representations on a 3D-rendered head surface as provided by E-field modeling software, such as SimNIBS ^72^ or ROAST ^73^. Besides the electrode position itself, it is also considered highly important to provide visual information (a photo or sufficiently detailed schematic figure) regarding the skin-electrode interface (item 1.4, average rating score: 4.09), i.e., which conducting medium was used (e.g., gel/paste or saline solution with sponges), how contact with the skin was ensured if the hair was in between, as well as measures taken to restrict the location of the contact medium to control the effective size of the stimulation surface and prevent short circuits. While the amount of conductive medium (volume of saline solution or thickness of the layer of electrode paste) was rated of medium importance (item 1.5, average rating score: 2.91), this information, together with the evenness of its distribution across the electrode surface, is relevant for the impedance as well as the current distribution in the skin (and potentially the brain). It can, however, be very difficult in practice to control this variable, given that gel is squeezed between electrode and head and saline solution flows away or evaporates, and it is thus helpful to also report potential countermeasures taken to control or measure this influence. In any case, electrode impedances should be measured directly before and after the experiment and be reported.

The introduction of any electrical wire into the MRI magnet bore may result in undesired artifacts and/or noise. Whereas the magnetic fields induced by the current in wires and electrodes during tDCS have been known to lead to false-positive activation in BOLD fMRI, tACS is far less prone to this artifact since the AC induces relatively rapid polarity switching magnetic fields that time average to zero net effect ^36^. However, any electrical cabling used in tES-MRI experiments may act as a transmitter of RF energy from outside the MRI shielded environment, and therefore may potentially increase electromagnetic RF interference with the MRI signal--even with the stimulator switched off. It is therefore extremely important to use an RF filtering method to suppress any external electromagnetic noise that may find its way into the scanner room using the stimulator’s cabling as a tunnel. Currently, there are two hardware configurations for addressing this issue ^29^: (1) The RF waveguide setup, which includes two filter boxes positioned outside and inside of the scanner room and cables running from the MRI control room through the RF waveguide tube ^37,38^; (2) The RF penetration panel setup, which includes an RF filter adapter connected directly to the RF penetration panel and MRI ground, positioned outside of the scanner room with cables running from the stimulator in the MRI control room through filter and RF penetration panel and to the electrode leads in the scanner room ^74^.

It is therefore recommended unanimously by the contributors to report this information which was rated to be moderately important (item 1.7, average rating score: 3.50) as precisely as possible. Additionally, it is recommended that the authors provide details regarding the attenuation characteristic of RF filtering (recommendation 1.7.1, recommended by 50% of the contributors) ^47,49^. For instance, “in the case of concurrent tDCS-fMRI, the characteristic bandwidth of the stop band of the filters on the DC path have been chosen to provide an approximate attenuation of 60 dB within a frequency range of 20–200 MHz to mitigate the radio frequency noise, protecting common strength MRI scanners such as 1.5T and 3T which operate at Larmor frequencies of approximately 64 MHz and 128 MHz during fMRI (proton imaging) ^47^”.

The wire routing pattern is also an important methodological detail when using transcranial stimulation simultaneously with fMRI measurement to increase replicability and validity of a study. However, this factor was rated as moderately important overall (item 1.8, average rating score: 3.43). It is important to make sure that the wires/cables do not create loops and run parallel to the bore axis as they approach and exit the scanner. It is also recommended by 58% of the contributors to ensure that after the subject enters the scanner, no loop can be created subsequent to entry due to wire movements, and practical measures to avoid them should be stated explicitly (recommendation 1.8.1). An example is a protocol reported by Williams and colleagues in which they emphasize that regarding stimulator setup, they ensured that no loop was made by the wire and it was placed along the wall of the room ^38^. It might be of importance (recommendation 1.8.2, recommended by 61% of the contributors) to include a figure illustrating the wiring details, such as the length of the cables required to connect inner and outer filter boxes, how the cables are connected to the electrodes, in which direction the cables are leaving the head, how multiple connecting cables are managed together, and depending on the geometry of the head coil, how the cables are entering the coil. Researchers are also encouraged (recommendation 1.8.3, recommended by 59% of the contributors) to report how they controlled cable motion inside the scanner (e.g., via sandbag, tape, etc.). One reason for doing so is to make sure that no loop is created by movements ^37^. The contributors stress the importance of reporting if there were any deviations from the device manufacturers’ recommendations due to study purposes (recommendation 1.8.4 recommended by 78% of the contributors). There are different institutional policies in various countries regarding the use of electrical stimulators during MR imaging (e.g., permission to transfer electrical current through the penetration panel); However, only 29% of responders recommended reporting limitations at the levels of institutions/countries based on regulations or policies (recommendation 1.8.5). This information might not be required as it does not affect the results of the study if the methods are transparent. The full potential of simultaneous tES and fMRI acquisition, such as dynamic monitoring of the brain during tES, can only be explored if the data of both systems are temporally synchronized. As the analysis depends critically on properly timed stimulation, it is crucial to synchronize imaging and stimulation. It is therefore recommended by the contributors to report this information which was rated to be moderately important (item 1.9, average rating score: 3.70) as precisely as possible. In general, to address this issue, the presentation computer receives a volume trigger TTL output from the MRI scanner and also sends output TTL triggers to the stimulator at desired stimulation times through a stimulus presentation software ^75^. Additionally, as synchronization protocols vary from center to center, it is recommended to clearly specify which method was used when sending the trigger pulse (recommendation 1.9.1, recommended by 96% of the contributors). There are several methods for addressing this issue, e.g., (1) USB, (2) parallel port, or (3) other additional devices. Two devices most commonly used for sending the trigger pulse include a USB DAQ device, which works well for the Psychtoolbox software package ^76^, and a USB-to-Serial port device, which works well for the E-Prime software package ^77^.

### Safety and Noise Tests

Reporting technical parameters that can be safety-relevant was considered as highly important (item 2.1, average rating score: 4.37). Ensuring the safety of the equipment for all possible MR environments and applications is usually not possible. Rather, most equipment is demonstrated to be *MR conditional*, i.e., safe under specific usage conditions in specific MR environments ^78^. This implies that the same equipment might still pose safety risks when used in untested scenarios, requiring a reevaluation of its safety.

Manufacturers of tES equipment should clearly document the safety-relevant technical parameters and settings used for their testing to ensure that users can replicate those appropriately. While it was less frequently recommended to repeat these parameters in the paper (recommendation 2.1.2, recommended by 51% of the contributors), deviations should be clearly reported, including the measures that were taken to ensure that safety was not compromised. To provide some guidance, the following paragraph gives a brief overview of aspects that can be safety-relevant and thus warrant consideration. Generally, external equipment brought inside the MR scanner might cause harmful effects via interaction with the *static magnetic field*, the *magnetic gradient fields*, and the *transmitted radiofrequency (RF) field* ^79^:

1. The *static magnetic field* exerts strong accelerating forces on ferromagnetic materials. In the case of tES-fMRI, using only non-magnetic materials for the cables and electrodes is a straightforward way that should be taken by the equipment manufacturer to prevent safety risks.
2. The *time-varying magnetic gradient fields* can create eddy currents in a conductive material that in turn result in mechanical forces via their interaction with the static field. This effect seems less relevant in the case of tES-fMRI for which the cables are the only high conductive parts. As they do not form closed high-conductive loops at the low electromagnetic frequencies corresponding to the time-varying gradient fields and are interrupted by the head, the electrodes, the stimulator, and often also safety resistors, the currents induced by gradient field switching are weak. This effect might, however, cause vibrations of the cables and might contribute to local nerve stimulation underneath the electrodes, while serious adverse effects such as burns due to tissue heating are unlikely ^79^.
3. Interactions of cables and electrodes with the *transmitted radiofrequency (RF) field* can potentially lead to local tissue heating and burns, which has been described, e.g., for electrocardiogram equipment ^80,81^. The MR scanner controls the transmit power to ensure that the specific absorption rate (SAR), i.e., the mean power deposition per unit tissue weight, stays within safe limits everywhere in the body. When cables are brought into the scanner, they can absorb and redistribute RF energy. By that, they might heat up and additionally locally focus RF energy in close-by body tissue. Both mechanisms can cause burns. They can occur for wire loops, but also for more or less straight cables that act as antennas, depending on several parameters including wire length and path, the terminal conditions at the electrodes, the frequency of the RF transmit field (linearly scaling with the MR field strength as long as only hydrogen nuclei are imaged), the spatial extent of the RF transmit field and the head and body position inside the field. Some of these parameters are difficult to standardize in practice so it is worth noting that the absence of heating in a test scan might not necessarily generalize. The safety of cables can be relevantly improved by adding resistors or cable traps or using lower conductive carbon instead of copper wires to systematically reduce or fully prevent the occurrence of standing waves. While these measures can be very effective, expert knowledge is required when implementing them to ensure that they work as intended and in a wide range of practical scenarios ^82^. When space allows, a simple measure to reduce the risk of burns is to ensure a physical distance between the cables and the skin. However, this does not help to prevent burns around points of high resistance, e.g., at the connection to the electrode, which is generally more likely.

The electrodes and gel are far less conductive than metal so that their interaction with the RF transmit field is relevantly smaller. However, as the rubber electrodes still have better ohmic conductance than body tissue (e.g., ∼30 S/m for the silicon rubber), they can cause a redistribution of the electric field that is created by the RF transmit field inside the head ^83^. This effect can change the local SAR distribution and potentially cause local skin heating. Its strength depends on the size, shape, and position of the electrodes, with the tendency that heating will be stronger for larger and thicker rubber electrodes.

The strength and duty cycle of the RF transmit field depends on the MR sequence type, which translates to the amount of local SAR increases that might occur due to electrodes or cables. Standard gradient-echo EPI used for functional brain imaging has comparatively low SAR. The SAR of newer multiband EPI and in particular turbo spin echo sequences (RARE, TSE, FSE, FLAIR, T2-SPACE, …) for T2-weighted structural imaging can be close to the allowed limits and might exceed these limits locally when cables and electrodes are present.

To summarize, interactions of the tES cables and electrodes with the RF transmit field depend on several parameters, which can make it difficult to generally ensure that local heating of the skin is kept within safe limits. Measures such as resistors added to the cables can reduce the risk of inducing adverse effects, but it remains important that the tES equipment is employed within the technical parameter ranges that are cleared by the manufacturer. These parameters include the MR field strength, the type of transmit coil (body coil vs birdcage coil or transmit array), the MR sequence type and settings, the cable paths, the electrode sizes as well as their shape, position, and material.

According to our knowledge, with the concurrent application methods, no higher number of reported adverse events (AEs) compared to conventional tES applications and no serious adverse events (SAEs) have been reported ^84^. Nevertheless, the study protocol must always comply with the safety standards for both tES and MRI and these parameters should be carefully documented in the protocol/paper. More detailed suggestions and recommendations of experts can be found in Table 2.

Experiments should always start with safety testing when a new protocol is applied. These safety tests should include, but are not limited to, impedance testing, temperature testing (any temperature change under electrodes) ^85^, and electric current tolerance testing (recommendation 2.2.1, recommended by 71% of the contributors). As suggested by at least 45 respondents (Figures 2 and 3), it is highly recommended to report impedance changes before and during the course of scanning and using a gel under the electrode (and not saline-soaked sponges) in order to avoid impedance increase (recommendation 2.5.1, recommended by 88% of the contributors).

The measurement of signal-to-noise ratio (SNR) was rated 3.91 (item 2.4), reflecting an important aspect in tES-fMRI studies. A small number of papers reported SNRs during the concurrent application of tES and fMRI, although it is well-known that electrical equipment can compromise image SNR via several mechanisms resulting in distorted images and false-positive changes ^85^. The stimulator is connected to the MR-compatible electrodes by specially designed leads. In some devices, the stimulating leads are passed through a waveguide tube in the MR cabin wall and through a radiofrequency filter module, consisting of two filter boxes. In other stimulators, there is only one filter attached to the patch (penetration) panel of the MRI (to ensure that the Faraday cage of the MRI room is not opened) and there is no noise induced during the normal MRI image acquisition. In spite of these safeguards, a small amount of noise is frequently present.

At least two papers reported susceptibility artifacts underneath the electrodes restricted to the skull layer with no visual evidence of any distortion in brain EPI images ^39,47^. Another study using fMRI measurements during tES in cadavers observed significant BOLD signal changes ^36^. Therefore, careful inspection of the SNR in different conditions during data acquisition is of critical importance to diminish errors and issues related to false-positive results. However, sometimes it is very difficult to deal with tES-fMRI artifacts because they might emerge sporadically, can be stimulation protocol and montage specific (e.g., tDCS seems to induce more noise than tACS) and often are not reproducible. Artifacts can be caused by many factors, by the noise of the stimulator itself, by the electrode/cable positions relative to the direction of the magnetic field, or by individual anatomical differences. Artifact removal is not trivial and may depend on the applied task in the scanner and processing methods. Beyond manual inspection, in a recent study, independent component analysis (ICA) was used to automatically remove noise in concurrent tDCS-fMRI ^86^. Manual inspection suggested that by applying this method, noise was successfully removed from the voxel’s time series.

As suggested in Table 2, our recommendations are:

1. tES manufacturers should state in the manual to what degree SNR changes during stimulation. As SNR will depend on the local settings, the type of the scanner (e.g., its shimming performance), and the MR sequences, several tests are suggested at different locations. This scan can be achieved using phantoms and in human subjects targeting different ROIs, tES-doses, and electrode positions ^47^. Basing the tests on the spherical agar phantom and the procedures outlined in the Function Biomedical Informatics Research Network (fBIRN) protocol would be a good starting point to ensure that the results of the quality tests are comparable between different MR sites and tES equipment ^87^. They should be complemented by measurements of the RF noise spectrum using the standard test sequences provided by scanner manufacturers, and by field mapping sequences to quantify the distortion of the static magnetic field induced by tES equipment ^88^.
2. When a new stimulator or protocol is tested, pilot in-scanner investigations, first using phantoms and later healthy human participants, are necessary and any incident or the absence of incidents should be reported.
3. SNR testing should always be done before the study starts (pilot measurements).
4. Later, during the study phase, when artifacts/SNR changes occur, it should be reported how many participants or runs were excluded from the analysis due to artifacts. Visualization of the artifacts is suggested.
5. If other devices are involved during the tES-fMRI session, it should be tested whether these devices or the interactions modify SNR. In the protocol, it should be clearly stated how tES-induced noise can be or was separated from other types of noise.

Subjective tolerance was reported in only 33% of the concurrent tES-fMRI articles in our systematic review. A gradual change in intolerance/side effects (itching sensation, burning, pain) may be the source of non-tES induced BOLD changes. This is particularly important for online tES studies but may also have an impact on offline tES. Subjective intolerance that leads to study discontinuation should always be reported. In addition, it is recommended that gradual subjective intolerance is reported (recommendation 2.3.1, recommended by 98% of the contributors). The Comfort Rating Questionnaire (CRQ) offers a good way to do this ^89^. It measures sensations such as pain, tingling, burning, fatigue, nervousness, concentration, vision, sleep disturbances, headaches, and flashes of light before, during and after stimulation, wherever possible as a visual analogue scale between 1 (not at all) - 10 (extreme). Subjective intolerance reporting (item 2.3) was rated 4.33 by the contributors. This indicates consensus that it is important to report this item in publications.

### Methodological Factors

It is crucial for studies to be precise about the timing of tES application relative to fMRI acquisition and also relative to any behavioral task performed, for both technical and experimental reasons. The checklist contains two items related to this specific point: items 3.1, “Concurrent tES-fMRI Timing” and 3.2, “Imaging Session Timing”. It is the committee’s position that these items should be reported with precise details (Table 3).

This is to address three issues in particular. First, tES-fMRI studies targeting sensorimotor cortex have clearly shown that the acute stimulation effects during tDCS are not the same as its post-stimulation effects ^6,7^. Therefore, knowledge about fMRI effects during stimulation cannot be simply extrapolated. Second, it is also increasingly recognized that brain state is an important determinant of the BOLD response to tES ^75,86^. This is not surprising, given that tES is thought to modulate spontaneous neuronal activity via subthreshold changes of membrane polarization without directly eliciting action potentials. Thus, it is vital that studies report exactly when stimulation was applied during the task, so that findings can be interpreted with knowledge of the underlying brain state. One final issue is that there is still relatively little known about the duration and nature of after-effects of tES. Early tDCS studies used the classical bipolar montage to stimulate the motor hand area and measured the motor-evoked potential (MEP), rather than fMRI, as the physiological outcome. These seminal studies suggest that at least three minutes of continuous stimulation are needed to produce after effects on corticomotor excitability ^21^ and that prolongation of stimulation within specific windows can prolong after-effects ^90,91^. But these dose-response relationships have been less frequently studied for other brain areas ^92,93^ and have yielded somewhat divergent results. This also applies to the concurrent tES-fMRI approach ^27^.

We recommend that the timeline of experimentation is reported in detail together with other design related information, such as counterbalancing of scans for within-subject studies, and whether subjects are repositioned in between scan runs should also be reported. This level of detail helps the reader to evaluate the results in the appropriate context and will enable the replication of techniques by other researchers.

For concurrent tES/fMRI studies, besides the importance of reporting the timing of tES relative to the timing of fMRI, it is also important to report the precise timing of tES *within* a particular fMRI imaging sequence (item 3.2, average rating score: 4.09). This is critical in order to assess the temporal relationship between tES and physiological activity acquired from fMRI, particularly in scenarios where the stimulation itself is dynamic (e.g., tACS, or during the ramping up/down of tDCS), which may then lead to different dynamics in fMRI-recorded physiological activity. For example, in a tDCS-fNIRS (functional near-infrared spectroscopy) study on a small sample of stroke patients, anodal tDCS resulted in non-stationary changes in blood oxygenation at the start of stimulation, possibly due to stimulation-induced changes in blood vessel dilation or neurovascular coupling ^94,95^. This issue deserves additional consideration in block designs where stimulation is applied in an on-off-on sequence. Here, physiological effects obtained by fMRI could possibly further be confounded by carryover or homeostatic effects due to repeated stimulation ^96,97^. Researchers interested in employing such a block design or repeated-stimulation approach may want to consider assessing the temporal stability of their stimulation protocol on the fMRI signal of interest. In all cases, the experts advise that care should be taken to report the precise stimulation start time in relation to the start of the imaging sequence, and a diagram or schematic be included along with the methodological description in order to provide maximum clarity to readers.

Reporting tES-associated sensations is crucial when using tES in any experimental or clinical setting, both for safety and methodological reasons. This item was rated with 4.06 which shows a high agreement within the panel regarding its relevance when reporting the methods in tES-fMRI studies. Different stimulation protocols can induce different sensory experiences and associated brain activity changes, which can in principle be confounded with true direct tES effects. Experimenters should consider this as a possible confound, e.g., when comparing between stimulation protocols and/or montages. For example, in the case of tACS, cutaneous sensation and phosphene perception, i.e., perceiving an illusionary flash-like light evoked by electric or magnetic pulses, are frequency-specific ^98^ and also differ between brain states (e.g., lighting conditions, eyes-open vs eyes-closed ^51,99^). Moreover, phosphene intensities have been shown to correlate with tACS-induced BOLD signal changes in the insular cortex, during 10Hz stimulation ^51^. In addition to phosphenes, or cutaneous sensations, different tES montages can potentially induce different levels of discomfort, especially while participants are lying in the MRI (e.g., depending on the distance between the electrodes and the RF coil and also on electrode location (e.g., if located on the back of the head)). When interpreting tES effects, it is important to carefully evaluate associated experiences to separate secondary from direct tES effects.

Reporting tES-associated sensory experiences is also crucial for safety reasons (see also section 4.2, subjective intolerance item). Asking participants to report on several factors such as electric current tolerance, headache, nausea, burning sensation, and pain can help experimenters to better monitor unwanted tES side effects, which will help to guarantee the safety of concurrent tES-fMRI protocols. Therefore, we recommend to assess and report tES-associated experiences before, during, and/or after tES (as appropriate). As stated in Table 3, we specifically recommend that: (1) tES associated sensory experience (e.g., tactile sensation, phosphene perception, burning sensation, and others) should be reported using rating scales or questionnaires (e.g., ^89,100^); additionally, participants should report whether they can differentiate between active and sham stimulation conditions (to assess the effectiveness of blinding whenever appropriate). The latter could be done by asking the participants to assign conditions in a forced choice manner. This would allow testing whether they perform above chance level in detecting real stimulation, even when consciously not being able to state a difference. (2) Electric current tolerance should be reported before entering the scanner room (if technically possible), inside the scanner, and/or before/during scanning (as appropriate). (3) Experimenters should report any instructions or additional training/tests that were conducted before the tES-fMRI session to make the experiment more suitable for the participant.

## Conclusion

The ContES checklist is a consensus-based product, which aims to promote best practices in reporting the relevant methodological details of concurrent tES-fMRI studies. We hope that the ContES checklist will encourage researchers to consider the scientific reasoning behind each methodological choice more thoroughly and report detailed methodological parameters of their studies more completely. This will improve the technical and scientific standard of concurrent tES-fMRI studies, and also help with the interpretability of the results and the reproduction of experiments. This checklist could also be useful when concurrent tES-fMRI study protocols are being designed and methodological parameters decided upon. Addressing the checklist in pre-registered protocols will enhance the scientific rigor and increase the replicability of protocols. As technological and methodological aspects of concurrent tES-fMRI studies diversify and the field advances over time, the steering committee of the checklist will work on future versions of the checklist to keep its details up-to-date. To ensure the feasibility of checklist application, we suggest considering reporting the “items” (Supplementary Table 6) as a “routine” requirement in concurrent tES-fMRI studies, and consideration of “additional recommendations” as “suggestions” to improve the methodological design and reporting of concurrent tES-fMRI studies (Supplementary Table 7). As with any checklist, the exact importance of each item will ultimately differ for each study, and it is the responsibility of the investigator, with support from regulatory and supervising bodies, to adapt the standards appropriately. It is impossible to anticipate every possible experimental set-up, equipment, or subject characteristic, and how these factors interact to influence important methodological and reporting considerations. Nonetheless, the development of generalized checklists provides standards and references for the research field, and a common language to discuss methodological and reporting concerns with a baseline framework. Ultimately, the impact of this checklist will depend on its use by authors, reviewers, and editors in the reporting, editing, and peer-review processes.

## Data Availability

The results of each phase were summarized and displayed on the OSF page: https://osf.io/f9j8z/

---

**Extended Data Fig. 1.**
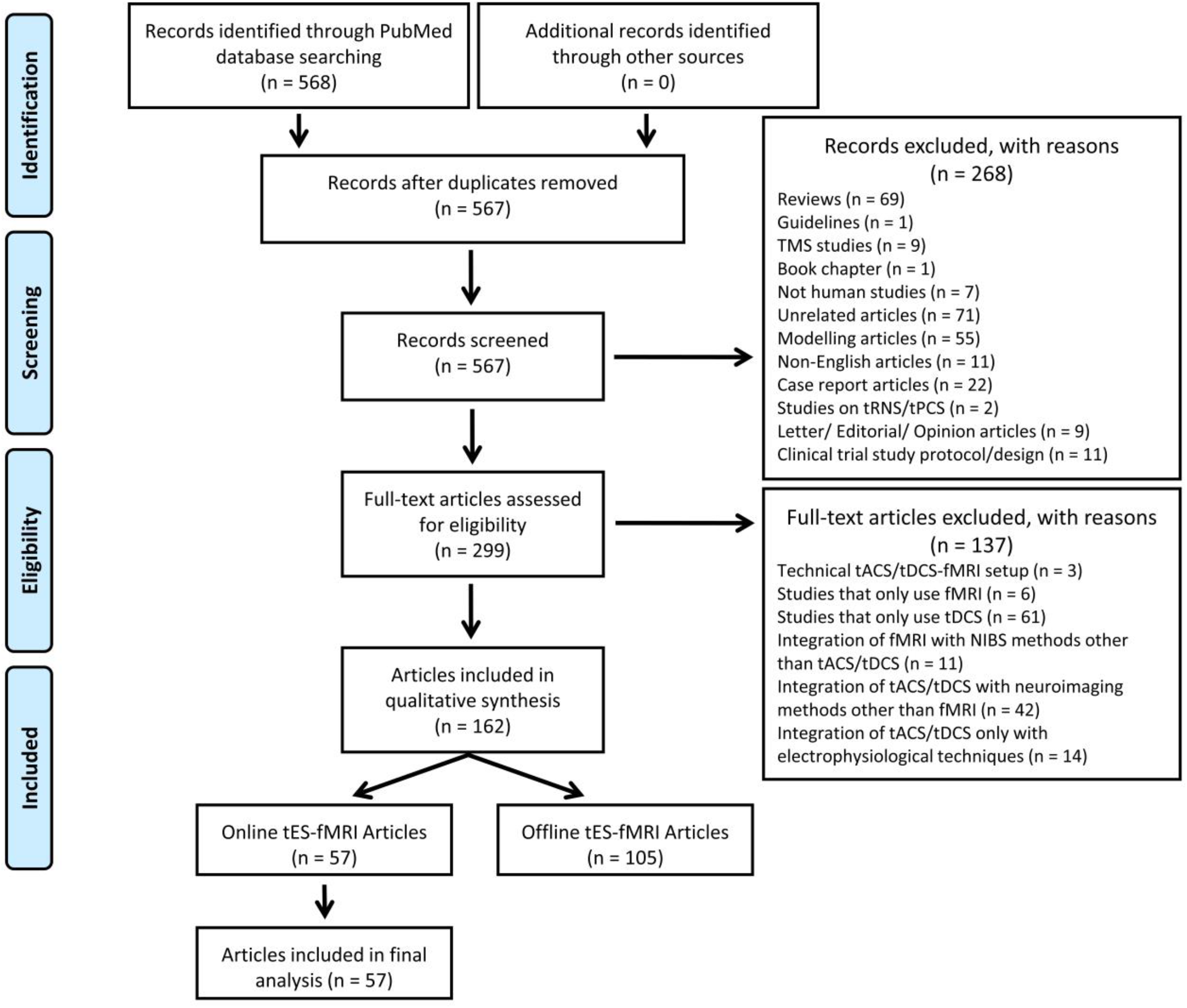
PRISMA flow diagram for concurrent tES-fMRI studies. Diagram of the literature search (identification) and selection process (screening, eligibility, inclusion).

**Extended Data Fig. 2.**
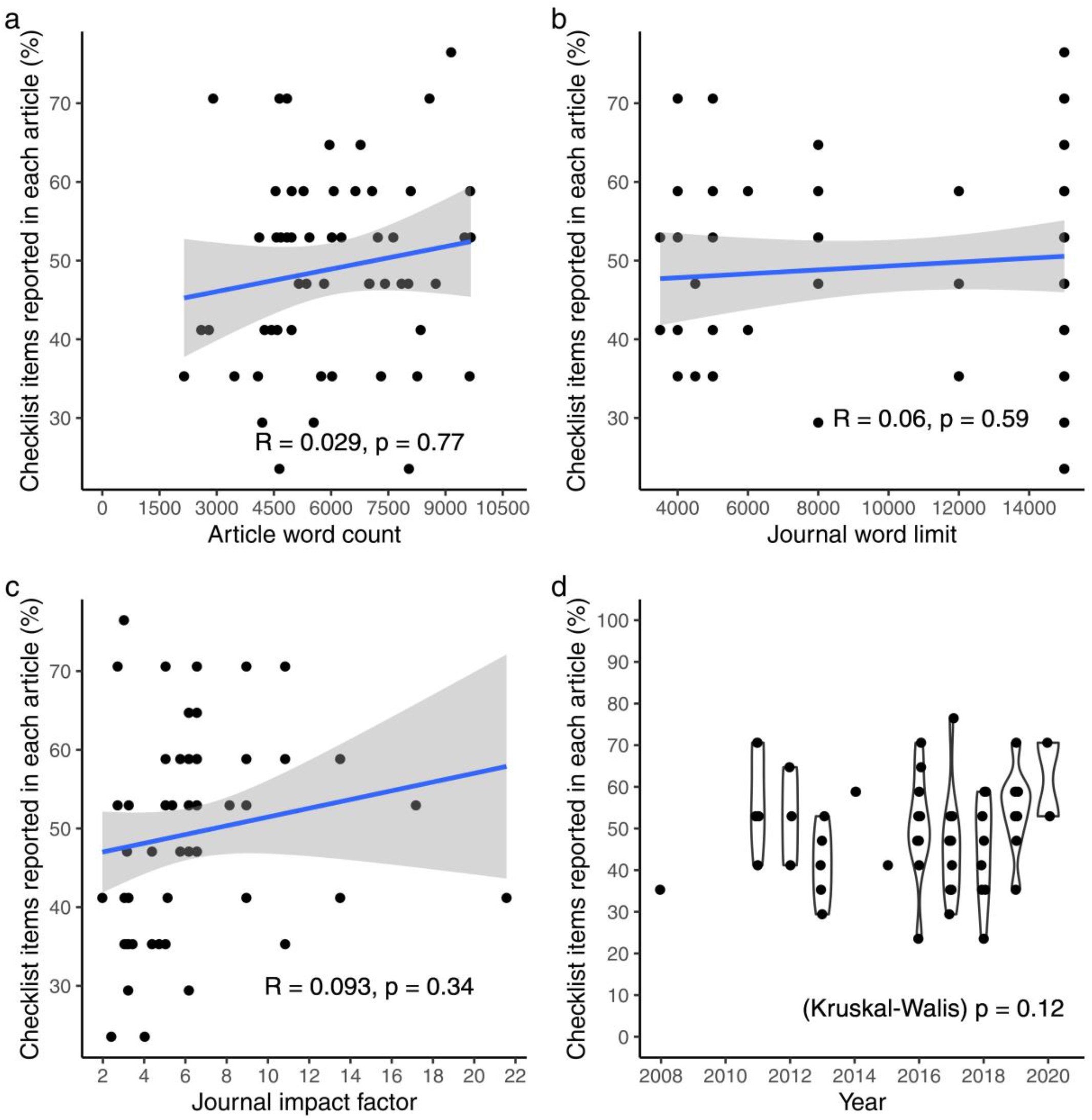
Relationships between reporting score and publication context. **a**, Relation between reporting score of each article with its word count (Note: Article word count is not exactly accurate, since it is measured by counting the words from the beginning of the introduction to the end of the discussion part, thus it might include the running title of each page, footnotes, and the captions of figures and tables). **b**, Relation between reporting score of each article with its journal word limit (Note: word limitation for journals with no word limitation is counted as 15000). **c**, Relation between reporting score of each article with journal impact factor. **d**, Article reporting scores across the years. Relationships of figure a, b, and c were assessed using linear regressions, while a Kruskal-Wallis test was performed for figure d.

## Funding Supports

Rany Abend is partially supported by the National Institute of Mental Health Intramural Research Program. Jorge Almeida is supported by the European Research Council (ERC) under the European Union’s Horizon 2020 research and innovation programme (Grant agreement No. 802553 - “ContentMAP”) and by grant PTDC/PSI-GER/30745/2017 from Fundação para a Ciência e a Tecnologia Portugal, and Programa COMPETE. Andrea Antal is supported by the Ministry for Science and Culture of Lower Saxony Germany (76251-12-7/19 ZN 3456) and by the BMBF (STIMCODE 01GP2124B). Daria Antonenko is supported by the Deutsche Forschungsgemeinschaft (DFG, German Research Foundation (AN 1103/3-1). Chris Baeken is supported by the Research Foundation – Flanders (FWO) and the Queen Elisabeth Medical Foundation for Neurosciences. Helen Barron is supported by the Medical Research Council (MRC) UK (MC_UU_00003/4 and MC_UU_12024/3). Til Ole Bergmann is supported by the Boehringer Ingelheim Foundation and the Deutsche Forschungsgemeinschaft (DFG, German Research Foundation, Grant 362546008). Marom Bikson is supported by grants from Harold Shames and the National Institutes of Health: NIH-NIDA UG3DA048502, NIH-NIGMS T34GM137858, NIH-NINDS 1R01NS112996, NIH-NINDS 1R01NS101362, NIH-NIMH 1R01MH111896, and NIH-NINDS 1R01NS095123. Matthew H. Davis is supported by the UK Medical Research Council (SUAG044/G101400). Hamed Ekhtiari is supported by the Laureate Institute for Brain Research (LIBR), Warren K. Family Foundation, Oklahoma Center for Advancement of Science and Technologies (OCAST, #HR18-139) and Brain and Behavior Foundation (NARSAD Young Investigator Award #27305). Valentina Fiori is supported by the Italian Ministry of Health (grant GR-2018-12365991). Peyman Ghobadi-Azbari is supported by the Cognitive Science and Technologies Council (grand CSTC-7761). Iman Ghodratitoostani is supported (grant number: 2013/07375-0) by Innovation and Diffusion of Mathematical Sciences Center Applied to Industry (CEPID-CeMEAI) of Sao Paulo Research Foundation (FAPESP), the University of Sao Paulo. Gadi Gilam is supported by the Redlich Pain Research Endowment and the Feldman Family Foundation Pain Research Fund. Gesa Hartwigsen is supported by the Max Planck Society and by the German Research Foundation (HA 6314/3-1, HA 6314/4-1 and HA 6314/9-1). Tobias U. Hauser is supported by a Sir Henry Dale Fellowship (211155/Z/18/Z) from Wellcome & Royal Society, an ERC Starting Grant, a grant from the Jacobs Foundation (2017-1261-04), the Medical Research Foundation, and a 2018 NARSAD Young Investigator grant (27023) from the Brain & Behavior Research Foundation. Christoph S. Herrmann is supported by the German Federal Ministry of Education and Research (BMBF, grant nrs. 16SV7787 and 13GW0273D) as well as the German Research Foundation under Germany’s Excellence Strategy (DFG, grant nr. EXC 2177/1 – Project ID 390895286). Chi-Hung Juan is supported by the Ministry of Science and Technology, Taiwan (grant no.108-2639-H-008-001-ASP; 108-2321-B-075 -004 -MY2) and sponsored by Taiwan Ministry of Education’s “Academic Strategic Alliance: Taiwan and Oxford University” project grant. Daniel Keeser is supported by the German Center for Brain Stimulation (GCBS) research consortium (Work Package 5, Grant no. 01EE1403E), funded by the German Federal Ministry of Education and Research (BMBF). Bart Krekelberg is supported by the National Institute of Neurological Disorders and Stroke and the National Institute of Mental Health under awards R01MH111766 and R21MH113917. Lucia M. Li is supported by an NIHR Clinical Lectureship, Academy of Medical Sciences Starter Grant and the NIHR Brain Injury MedTech Cooperative. Sook-Lei Liew is supported by the National Institutes of Health (K01 HD091283, R01 NS115845). Timothy J. Meeker is supported by a Postdoctoral Scholarship from the Neurosurgery Pain Research Institute at the Johns Hopkins Medical Institute and National Institutes of Health (R01 NS107602). Marcus Meinzer is supported by the German Research Foundation (DFG ME 3161/3-1). Michael A. Nitsche is supported by Deutsche Forschungsgemeinschaft (DFG) - Projektnummer 316803389 – SFB 1280 “Funded by the Deutsche Forschungsgemeinschaft (DFG, German Research Foundation) - Projektnummer 316803389 - SFB 1280, the German Federal Ministry of Education and Research (BMBF, GCBS grant 01EE1501), and the EU (NEUROTWIN, grant 101017716). Alexander Opitz is supported by RF1MH117428 and RF1MH124909. Christian Ruff is supported by an ERC Consolidator grant (BRAINCODES, Grant #725355) and by the Swiss National Science Foundation (Grant # 100019L_173248). Michaela Ruttorf was supported by Deutscher Akademischer Austauschdienst (DAAD - Project-ID 57212180). Gottfried Schlaug acknowledges support by U01NS102353 and R01MH111874. A. Duke Shereen is supported by National Institute of Neurological Disorders and Stroke under award R21NS115018. Hartwig R. Siebner holds a 5-year professorship in precision medicine at the Faculty of Health Sciences and Medicine, University of Copenhagen which is sponsored by the Lundbeck Foundation (Grant Nr. R186-2015-2138). Hartwig R. Siebner has received a Lundbeckfonden “Collaborative Alliance” grant (Grant Nr. R336-2020-1035). Hartwig R. Siebner and Axel Thielscher are supported by the Innovationfund Denmark - grant agreement number 9068-00025B. Charlotte Stagg holds a Sir Henry Dale Fellowship, funded by the Wellcome Trust and the Royal Society (102584/Z/13/Z). Benjamin Thompson is supported by CIHR grant 390283, CFI grant 34095 and NSERC grants RPIN-05394 and RGPAS-477166. Axel Thielscher was supported by the Lundbeck foundation (R244-2017-196 and R313-2019-622). Dagmar Timmann is supported by the Deutsche Forschungsgemeinschaft (DFG, German Research Foundation) - Projektnummer 316803389 - SFB 1280. Ines Violante and Tibor Auer are supported by the BBSRC (BB/S008314/1). Benedikt Zoefel is supported by the European Union’s Horizon 2020 research and innovation programme under the Marie Sklodowska-Curie grant agreement number 743482. The views presented in this manuscript represent those of the authors and not necessarily those of the funding agencies.

