## Supplemental Tables1-5 for "A Checklist for Assessing the Methodological Quality of Concurrent tES-fMRI Studies (ContES Checklist): A Consensus Study and Statement"

**Supplementary Table 1 | Supporting Evidence and Example Articles for each Checklist Item.** The "Supporting Evidence" column includes studies which demonstrate how each checklist item might affect the results of a concurrent tES-fMRI study and its importance for interpretability and generalizability. Where empirical evidence is scarce, results from adjacent fields in cognitive neuroscience, qualitative reviews, and the statement by the Committee on Best Practice in Data Analysis and Sharing are cited. The "Reporting Example" column includes a number of exemplar papers which have reported each item.

| Section/Topic | Item No | Main Items to Report | Supporting Evidence | Reporting Example |
|---|---|---|---|---|
| ***Technological Factors*** | | | | |
| **Manufacturer of MR Conditional Stimulator** | 1.1 | The brand and model (if a brand is providing different MR conditional models) for the MR conditional stimulator. | 1 | 2,3 |
| **MR Conditional Electrode Details** | 1.2 | The MR conditional electrode type (i.e., conductive polymer with or without a sponge or other conductive medium holders). | 4 | 5,6 |
| **Electrode Positioning** | 1.3 | The method for electrode placement over the head inside the scanner (i.e., targeting software, 10-20 convention with or without EEG cap, functional targeting (fMRI), computational head models or others). | 7–10 | 11,12 |
| **MR Conditional Skin-Electrode Interface** | 1.4 | The MR conditional skin-electrode interface (saline solution, conductive paste, gel, etc.). | 1,4,13 | 14,15 |
| **Amount of Contact Medium (Paste/Gel/Electrolyte)** | 1.5 | The amount or thickness of medium that is used for each electrode or a method to control this confounding variable. | 1,13,16 | 17,18 |
| **Electrode Placement Visualization** | 1.6 | Any photo/diagram/figure to precisely visualize the electrode montage inside the scanner and make replication possible. | 19–21 | 22,23 |
| **RF Filter** | 1.7 | The RF filtering method (stimulator device connected to the subject via penetration panel (e.g., RF filters from different brands) or connected via waveguide with RF boxes on either end). | 20 | 24,25 |
| **Wire Routing Pattern** | 1.8 | Wire routing pattern (out back of bore and around the control room or straight down front of bore to control room). | 20,26 | 27,28 |
| **tES-fMRI Machine Synchronization/Communication** | 1.9 | The synchronization/communication method between the tES device, the stimulus delivery PC, and the scanner. | 29 | 30,31 |
| ***Safety and Noise Tests*** | | | | |
| **MR Conditionality Specifics for tES Setting** | 2.1 | The technical specifications of the MR scanner, the applied fMRI sequences, and the used tES settings and configuration to fall within the specifics of MR conditionality based on tES manufacturer guideline. | 32,33 | 34,35 |
| **tES-fMRI Setting Test - Safety Testing** | 2.2 | The safety of the tES-fMRI setting. | 36–38 | 39,40 |
| **tES-fMRI Setting Test - Subjective Intolerance Reporting** | 2.3 | The number of cases that have not tolerated the tES/fMRI session (even if it is zero). | 41 | 42,43 |
| **tES-fMRI Setting Test - Noise/Artifact** | 2.4 | The noise/artifact induced by the tES setting in the fMRI signal with real human subjects or phantoms before starting the study (It can be reported or referred to previous studies with the same setting). | 19,44 | 40,45 |
| **Impedance Testing** | 2.5 | Impedance monitoring (i.e., before entering the scanner room and/or in the scanner room and/or inside the scanner and/or during scanning). | 46,47 | 48,49 |
| ***Methodological Factors*** | | | | |
| **Concurrent tES-fMRI Timing** | 3.1 | The timing of concurrent tES within the fMRI paradigm. | 29,50–52 | 53,54 |
| **Imaging Session Timing** | 3.2 | The imaging events before and after concurrent tES-fMRI and respective sequences. | 55,56 | 57,58 |
| **tES Experience Report** | 3.3 | The assessment of the subjective experience of receiving tES inside the scanner. | 41,59 | 60,61 |

**Supplementary Table 2 | Summary of concurrent tES-fMRI studies.** This table provides details about the 57 concurrent tES-fMRI studies, which were included in our database during developing the checklist. This table summarizes the number of recruited participants, the electrode montage, the intensity, duration, and type of stimulation.

| Author, Year | Participants | tES Montage | Intensity (mA) | Duration (min) | tES Method | Reference |
|---|---|---|---|---|---|---|
| (Hauser et al. 2016) | 48 Healthy subjects | Between CP5 and P5/Between Fpz and AF8 | 1 | 30 | tDCS | 2 |
| (Callan et al. 2016) | 28 Healthy subjects | P4/Left shoulder | 1 | 30 | tDCS | 62 |
| (Alekseichuk et al. 2016) | 16 Healthy subjects | Oz/Cz | 1 (peak-to-baseline) | 10 | tDCS, tACS | 3 |
| (Worsching et al. 2017) | 20 Healthy subjects | F3/F4 | 2 | 20 | tDCS | 5 |
| (Antonenko et al. 2017) | 48 Older adults | C3/Fp2 | 1 | 15 | tDCS | 53 |
| (Lin et al. 2017) | 18 Healthy subjects | F3/FP2 | 1 | 20 | tDCS | 6 |
| (Zheng et al. 2016) | 9 Healthy subjects | 1/3 of the distance between F8 and C6/Fp1 | 1 | 10 | tDCS | 11 |
| (Yang et al. 2017) | 32 Smokers | F3/F4 | 1 | 30 | tDCS | 12 |
| (Darkow et al. 2017) | 16 post-stroke aphasias | C3/Fp2 | 1 | 20 | tDCS | 54 |
| (Sotnikova et al. 2017) | 16 ADHD | F3/Cz | 1 | 20 | tDCS | 63 |
| (Lindenberg et al. 2016) | 24 Healthy subjects | C3/C4<br>C3/Fp2 | 1 | 30 | tDCS | 14 |
| (Holland et al. 2016) | 10 Healthy subjects | FC5/Fp2 | 2 | 20 | tDCS | 64 |
| (Barron et al. 2016) | 53 Healthy subjects | T6/Fp1 | 1 | 20 | tDCS | 15 |
| (Meinzer et al. 2015) | 18 MCI | Left ventral IFG/Fp2 | 1 | 20 | tDCS | 65 |
| (Meinzer et al. 2014) | 18 Older adults | C3/C4<br>C3/Fp2 | 1 | 30 | tDCS | 22 |
| (Antal et al. 2014) | 60 Healthy subjects | F2-Fpz/O2-P4<br>O2-P4/F2-Fpz | 1 | 20 | tDCS | 66 |
| (Martin et al. 2017) | 24 Healthy subjects | C3/C4<br>C3/Fp2 | 1 | 30 | tDCS | 67 |
| (Orlov et al. 2017) | 49 Schizophrenia | F3/Fp2 | 2 | 30 | tDCS | 57 |
| (Meinzer et al. 2013) | 20 Older adults | Left ventral IFG/Fp2 | 1 | 20 | tDCS | 23 |
| (Stagg et al. 2013) | 24 Healthy subjects | F3/Fp2<br>Fp2/F3 | 1 | 20 | tDCS | 58 |
| (Lindenberg et al. 2013) | 20 Healthy subjects | C3/C4<br>C3/Fp2 | 1 | 30 | tDCS | 24 |
| (Sehm et al. 2013) | 12 Healthy subjects | C4/Fp1<br>C4/C3 | 1 | 20 | tDCS | 50 |
| (Saiote et al. 2013) | 52 Healthy subjects | C3/Fp2<br>Fp2/C3 | 1 | 10 | tDCS | 68 |
| (Sehm et al. 2012) | 12 Healthy subjects | C4/Fp1<br>C4/C3 | 1 | 20 | tDCS | 51 |
| (Kwon and Jang 2012) | 9 Healthy subjects | C3/C4<br>C3/Fp2 | 1 | 2 | tDCS | 69 |
| (Meinzer et al. 2012) | 20 Healthy subjects | Left IFG/Fp2 | 1 | 17 | tDCS | 70 |
| (Zheng, Alsop, and Schlaug 2011) | 14 Healthy subjects | C4/FP1<br>Fp1/C4 | 1.4 | 3 ON and 4 OFF blocks during 10 min | tDCS | 71 |
| (Holland et al. 2011) | 10 Older adults | FC5/Fp2 | 2 | 20 | tDCS | 72 |
| (Alon et al. 2011) | 5 Healthy subjects | C4/Fp1 | 2 | 12.8 | tDCS | 25 |
| (Kwon and Jang 2011) | 12 Healthy subjects | C3/Fp2 | 1 | 2 | tDCS | 73 |
| (Antal et al. 2011) | 20 Healthy subjects | Between O1-P3/Between O2-P4 | 1 | 0.33 | tDCS | 74 |

| | | | | | | |
|---|---|---|---|---|---|---|
| | | Between O2-P4/Between O1-P3 | | | | |
| (Kwon et al. 2008) | 11 Healthy subjects | C3/Fp2 | 1 | 1.4 | tDCS | 75 |
| (Li, Violante, Leech, Hampshire, et al. 2019) | 26 Healthy subjects | F8/Right shoulder<br>Right shoulder/F8 | 2 | 3 blocks of anodal/cathodal /sham (each one 4.2 min) | tDCS | 48 |
| (Li, Violante, Leech, Ross, et al. 2019) | 26 Healthy subjects | F8/Right shoulder<br>Right shoulder/F8 | 2 | 3 blocks of anodal/cathodal /sham (each one 4.2 min) | tDCS | 52 |
| (Gilam et al. 2018) | 25 Healthy subjects | Fpz/Right shoulder | 1.5 | 22 | tDCS | 76 |
| (Falcone et al. 2018) | 28 Healthy subjects | P4/Left shoulder | 1 | 30 | tDCS | 77 |
| (Worsching et al. 2018) | 32 Healthy subjects | F3/F4<br>F3/Fp2<br>F4/F3 | 2 | 20 | tDCS | 49 |
| (Abend et al. 2018) | 19 Healthy subjects | Fpz/Oz | 1.5 | 20 | tDCS | 78 |
| (Antonenko et al. 2018) | 30 Older adults, 30 Young subjects | C3/Fp2 | 1 | 15 | tDCS | 27 |
| (Ulrich et al. 2018) | 22 Healthy subjects | Fpz/Right shoulder | 1.5 | 30 | tDCS | 79 |
| (Fiori et al. 2018) | 28 Healthy subjects | FC5/Fp2 | 1 | 24 | tDCS | 80 |
| (Vosskuhl, Huster, and Herrmann 2016) | 29 Healthy subjects | Oz/Cz | 0.1, 0.6, 0.2 (peak-to-peak) | 18 | tACS | 60 |
| (Cabral-Calderin et al. 2016) | 13 Healthy subjects | Oz/Cz | 1.5 (peak-to-peak) | 0.5 min ON and 0.5 min OFF during 7 min | tACS | 28 |
| (Bachinger et al. 2017) | 20 Healthy subjects | C3+C4/Oz | 1.5 (peak-to-peak) | 14 | tACS | 30 |
| (Chai et al. 2018) | 11 Healthy subjects | Oz/Cz | 1 (peak-to-peak) | 12.5 | tACS | 31 |
| (Weinrich et al. 2017) | 12 Healthy subjects | C3/Fp2 | 1 | 4 runs each 1.3 min | tACS | 81 |
| (Moisa et al. 2016) | 20 Healthy subjects | C3/Left shoulder | 1 (peak-to-peak) | 6 runs each 0.3 min | tACS | 44 |
| (Violante et al. 2017) | 20 Healthy subjects | F4+P4/T8 | 1 (peak-to-peak) | 11.6 | tACS | 29 |
| (Zoefel, Archer-Boyd, and Davis 2018) | 17 Healthy subjects | T7/C3 | 1.7 (peak-to-peak) | 30 | tACS | 34 |
| (Kim et al. 2019) | 12 Schizophrenia | P4/P3<br>F4/F3 | 2 | 20 | tDCS | 35 |
| (Küper et al. 2019) | 51 Healthy subjects | Active: 3cm laterally to the inion/Return: right buccinator muscle | 1.8 | 20 | tDCS | 42 |
| (Nissim et al. 2019) | 16 Older adults | F4/F3 | 2 | 12 | tDCS | 17 |
| (Antonenko et al. 2019) | 24 Healthy subjects | C3/Fp2<br>Fp2/C3 | 1 | 15 | tDCS | 18 |
| (Jamil et al. 2020) | 29 Healthy subjects | Left motor cortex hotspot/Right SO | 0.5, 1, 1.5, 2, sham | 15 | tDCS | 39 |
| (Kar et al. 2020) | 10 Healthy subjects | Between PO7-P3/Cz | 1 (peak-to-peak) | 8 | tACS | 40 |
| (Lefebvre et al. 2019) | 46 Healthy subjects | Montage1: HD over motor hotspot<br>Montage2: HD over left premotor cortex | 1 | 7 | tDCS | 82 |
| (Li, Violante, Zimmerman, et al. 2019) | 35 TBI | F8/Right shoulder<br>Right shoulder/F8 | 2 | 3 blocks of anodal/cathodal /sham (each one 4.2 min) | tDCS | 61 |

**Abbreviation:** ADHD: attention deficit hyperactivity disorder; MCI: mild cognitive impairment; SO: supraorbital; TBI: traumatic brain injury.

**Supplementary Table 3 | Characteristics of steering committee (SC) and expert panel (EP) members.**

| Demographic Variables | Frequency (%) | |
|---|---|---|
| | Steering Committee | Expert Panel |
| **Gender** | | |
| Male | 8 (62%) | 29 (59%) |
| Female | 4 (31%) | 11 (22%) |
| Other | 1 (8%) | 5 (10%) |
| No response | 0 (0%) | 4 (8%) |
| **Age (years)** | | |
| 30–40 | 4 (31%) | 23 (47%) |
| 40–50 | 6 (46%) | 10 (20%) |
| ≥50 | 3 (23%) | 10 (20%) |
| No response | 0 (0%) | 6 (12%) |
| **Highest Academic Degree** | | |
| Master of Science | 0 (0%) | 3 (6%) |
| Doctor of Medicine | 2 (15%) | 2 (4%) |
| Doctor of Philosophy | 9 (69%) | 37 (76%) |
| Doctor of Medicine/Philosophy | 2 (15%) | 3 (6%) |
| No response | 0 (0%) | 4 (8%) |
| **Country of Residence** | | |
| Austria | 0 (0%) | 1 (2%) |
| Belgium | 0 (0%) | 1 (2%) |
| Brazil | 0 (0%) | 1 (2%) |
| Canada | 0 (0%) | 1 (2%) |
| China | 0 (0%) | 1 (2%) |
| Denmark | 2 (15%) | 1 (2%) |
| France | 0 (0%) | 1 (2%) |
| Germany | 3 (23%) | 14 (29%) |
| Italy | 0 (0%) | 3 (6%) |
| Portugal | 1 (8%) | 0 (0%) |
| Switzerland | 0 (0%) | 3 (6%) |
| Taiwan | 0 (0%) | 1 (2%) |
| United Kingdom | 3 (23%) | 5 (10%) |
| United States | 4 (31%) | 12 (24%) |
| No response | 0 (0%) | 4 (8%) |
| **Primary Field of Research** | | |
| Cognitive Science | 1 (8%) | 8 (16%) |
| Neuroscience | 11 (84%) | 24 (49%) |
| Psychiatry | 0 (0%) | 5 (10%) |
| Psychology | 0 (0%) | 5 (10%) |
| Others | 1 (8%) | 3 (6%) |

| | | |
|---|---|---|
| No response | 0 (0%) | 4 (8%) |
| **Primary Place of Work** | | |
| Business/Industry | 0 (0%) | 3 (6%) |
| Hospital | 1 (8%) | 9 (18%) |
| Independent Research Institute | 2 (15%) | 3 (6%) |
| University | 9 (69%) | 29 (59%) |
| Hospital and University | 1 (8%) | 0 (0%) |
| Others | 0 (0%) | 1 (2%) |
| No response | 0 (0%) | 4 (8%) |
| **Length of Time Spent in tES or fMRI Research (Years)** | | |
| Less than 5 | 0 (0%) | 1 (2%) |
| 5–10 | 3 (23%) | 10 (20%) |
| 10–20 | 5 (38%) | 25 (51%) |
| ≥20 | 4 (31%) | 9 (18%) |
| No response | 1 (8%) | 4 (8%) |
| **Length of Time Spent in tES-fMRI Research (Years)** | | |
| Less than 5 | 2 (15%) | 16 (33%) |
| 5–10 | 7 (54%) | 23 (47%) |
| 10–20 | 2 (15%) | 4 (8%) |
| ≥20 | 1 (8%) | 0 (0%) |
| No response | 1 (8%) | 6 (12%) |

**Supplementary Table 4 | Concurrent tES-fMRI (ContES 2021) checklist, short form.**

The ContES checklist is designed to provide a short list of the main items that every concurrent tES-fMRI study should consider in the final report/paper. These items are designed as simple questions to appraise articles with Yes or No answers. Authors could provide a filled checklist including the line/page where the item is addressed in the manuscript as a supplement in the process of manuscript submission for peer reviewed journals. Additionally, the checklist provides a list of recommendations for each item that could increase the quality of reporting. Although the checklist is designed primarily to guide the development of research reports, the items and recommendations can be considered when concurrent tES-fMRI studies are being designed as well.

| Section/topic | Item No | Main Items to Report | Page/Line |
|---|---|---|---|
| ***Technological Factors*** | | | |
| **Manufacturer of MR Conditional Stimulator** | 1.1 | The brand and model (if a brand is providing different MR conditional models) for the MR conditional stimulator. | |
| **MR Conditional Electrode Details** | 1.2 | The MR conditional electrode type (i.e., conductive polymer with or without a sponge or other conductive medium holders). | |
| **Electrode Positioning** | 1.3 | The method for electrode placement over the head inside the scanner (i.e., targeting software, 10-20 convention with or without EEG cap, functional targeting (fMRI), computational head models or others). | |
| **MR Conditional Skin-Electrode Interface** | 1.4 | The MR conditional skin-electrode interface (saline solution, conductive paste, gel, etc.). | |
| **Amount of Contact Medium (Paste/Gel/Electrolyte)** | 1.5 | The amount or thickness of medium that is used for each electrode or a method to control this confounding variable. | |
| **Electrode Placement Visualization** | 1.6 | Any photo/diagram/figure to precisely visualize the electrode montage inside the scanner and make replication possible. | |
| **RF Filter** | 1.7 | The RF filtering method (stimulator device connected to the subject via penetration panel (e.g., RF filters from different brands) or connected via waveguide with RF boxes on either end). | |
| **Wire Routing Pattern** | 1.8 | Wire routing pattern (out back of bore and around the control room or straight down front of bore to control room). | |
| **tES-fMRI Machine Synchronization/Communication** | 1.9 | The synchronization/communication method between the tES device, the stimulus delivery PC, and the scanner. | |
| ***Safety and Noise Tests*** | | | |
| **MR Conditionality Specifics for tES Setting** | 2.1 | The technical specifications of the MR scanner, the applied fMRI sequences, and the used tES settings and configuration to fall within the specifics of MR conditionality based on tES manufacturer guideline. | |
| **tES-fMRI Setting Test - Safety Testing** | 2.2 | The safety of the tES-fMRI setting. | |
| **tES-fMRI Setting Test - Subjective Intolerance Reporting** | 2.3 | The number of cases that have not tolerated the tES/fMRI session (even if it is zero). | |
| **tES-fMRI Setting Test - Noise/Artifact** | 2.4 | The noise/artifact induced by the tES setting in the fMRI signal with real human subjects or phantoms before starting the study (It can be reported or referred to previous studies with the same setting). | |
| **Impedance Testing** | 2.5 | Impedance monitoring (i.e. before entering the scanner room and/or in the scanner room and/or inside the scanner and/or during scanning). | |
| ***Methodological Factors*** | | | |
| **Concurrent tES-fMRI Timing** | 3.1 | The timing of concurrent tES within the fMRI paradigm. | |
| **Imaging Session Timing** | 3.2 | The imaging events before and after concurrent tES-fMRI and respective sequences. | |
| **tES Experience Report** | 3.3 | The assessment of the subjective experience of receiving tES inside the scanner. | |

*We strongly recommend that this checklist be read in conjunction with the ContES checklist development and consensus paper. The paper should be cited when using the checklist as well.

**Supplementary Table 5 | Concurrent tES-fMRI (ContES 2021) checklist, long form.**

The ContES checklist is designed to provide a short list of the main items that every concurrent tES-fMRI study should consider in the final report/paper. These items are designed as simple questions to appraise articles with Yes or No answers. Authors could provide a filled checklist including the line/page where the item is addressed in the manuscript as a supplement in the process of manuscript submission for peer reviewed journals. Additionally, the checklist provides a list of recommendations for each item that could increase the quality of reporting. Although the checklist is designed primarily to guide the development of research reports, the items and recommendations can be considered when concurrent tES-fMRI studies are being designed as well.

| Section/topic | Main Items to Report | Page/Line | Additional Recommendations |
|---|---|---|---|
| ***Technological Factors*** | | | |
| **1.1. Manufacturer of MR Conditional Stimulator** | The brand and model (if a brand is providing different MR conditional models) for the MR conditional stimulator. | | |
| **1.2. MR Conditional Electrode Details** | The MR conditional electrode type (i.e., conductive polymer with or without a sponge or other conductive medium holders). | | 1.2.1. Report conductive properties of the MR conditional electrodes, cables, contact medium, and other conductive elements, including the position and materials used for the electrode-cable connections (Saturnino et al., 2015). This is especially important if they are not from an established manufacturer or not well described in the prior literature. However, even for well-established equipment, these details are critical to report to ensure replicability. |
| **1.3. Electrode Positioning** | The method for electrode placement over the head inside the scanner (i.e., targeting software, 10-20 convention with or without EEG cap, functional targeting (fMRI), computational head models or others). | | 1.3.1. Report electrode positioning as precisely as possible to facilitate reproduction. It is usually inadequate to simply report an anatomical target, for example, "the anodal electrode was placed over M1". |
| | | | 1.3.2. Report whether electrode positioning is based on the individual anatomy or a group template if imaging or head modeling is used for electrode positioning. |
| | | | 1.3.3. Report how electrode positioning is performed at the individual participant level. For example, was a neuronavigation system used or the EEG 10-20 system or something else. |
| | | | 1.3.4. Report the methods to ensure that the same electrode locations were used again if there are multiple sessions. |
| | | | 1.3.5. Report clearly how the electrodes are held in place inside the scanner including use of head-gear or customized supports. |
| | | | 1.3.6. Report how electrodes and their connecting cables over the head are located in relationship to the MR head coil while the subject is laying down inside the scanner and how the head was held in place - e.g., pillows, foam, etc. to ensure that position of head/electrodes remain in the same place during the scans while the convenience of the participant is ensured. |
| | | | 1.3.7. Report a post-hoc validation of the electrode positioning based on anatomical images with the electrodes in place if practical. For optimal validation, current density models based on anatomical images may be used (e.g., ROAST, SIMNIBS, etc.). It would be even better to directly measure the electric fields using magnetic resonance current density imaging (MRCDI) and MR electrical impedance tomography (MREIT) (Göksu et al., 2018), however, MREIT and MRCDI are still not available in most of the institutes. |
| **1.4. MR Conditional Skin-Electrode Interface** | The MR conditional skin-electrode interface (saline solution, conductive paste, gel, etc.). | | 1.4.1. Report a photo or a schematic figure or technical details showing in a reproducible way how the electrode with the MR conditional skin-electrode interface is connected to the cranium (including a view from the underneath of the electrode if needed). If headgear or headstraps obscure the electrodes, you may provide an image without the headstraps. |
| | | | 1.4.2. Report any other MR-specific strategies to restrict the contact medium (such as within an electrode holder) to avoid short circuits. |
| **1.5. Amount of Contact Medium (Paste/Gel/Electrolyte)** | The amount or thickness of medium that is used for each electrode or a method to control this confounding variable. | | 1.5.1. Report technical details/difficulties in measuring the thickness of the layer of conductive material underneath the electrodes and how cream/gel underneath the electrodes is evenly distributed. Although this can be important, mainly when having big electrodes, in practice, the amount of cream/gel underneath the electrodes may not be evenly distributed. Developing new methods to measure, control, and report this important variable are desired. Reporting the impedance (before, during, and after stimulation) provides insight on electrode contact quality, but is not in itself a substitute for controlling and reporting contact medium parameters. |
| **1.6. Electrode Placement Visualization** | Any photo/diagram/figure to precisely visualize the electrode montage inside the scanner and make replication possible. | | |
| **1.7. RF Filter** | The RF filtering method (stimulator device connected to the subject via penetration panel (e.g., RF filters from different brands) or connected via waveguide with RF boxes on either end). | | 1.7.1. Report the attenuation characteristic of the RF filtering. |
| | | | 1.7.2. Report any potential regulatory consideration/limitation at the institute/university/country level. |
| **1.8. Wire Routing Pattern** | Wire routing pattern (out back of bore and around the control room or straight down front of bore to control room). | | 1.8.1. Report whether/how the state of the cables is checked after the subject entering the scanner to avoid creating any loops. |
| | | | 1.8.2. Report the length of the cables required to connect inner with outer box using box cable, how the cables are connected to the electrodes, in which direction the cables are leaving the head, how multiple connecting cables are managed together, and depending on the geometry of the head coil, how the cables are entered into the coil. A sketch might be helpful to visualize these details. |
| | | | 1.8.3 Report how the cables and filter boxes are secured to prevent motion during the scan (i.e., sandbag, tape, etc.). |
| | | | 1.8.4. Report if there are any modifications from manufacturer recommendations. |
| | | | 1.8.5. Report any potential regulatory consideration/limitation at the institute/university/country level. |
| **1.9. tES-fMRI Machine Synchronization/Communication** | The synchronization/communication method between the tES device, the stimulus delivery PC, and the scanner. | | 1.9.1. Report any synchronization between tES and MRI. Synchronization/communication can be TTL scanner sync pulse to trigger/sync (tES and/or non-tES) stimulus recorded via USB/parallel port/NI device; use of markers for tES, or manual triggering of the TES device. |
| ***Safety and Noise Tests*** | | | |
| **2.1. MR Conditionality Specifics for tES Setting** | The technical specifications of the MR scanner, the applied fMRI sequences, and the used tES settings and configuration to fall within the specifics of MR conditionality based on tES manufacturer guideline. | | 2.1.1. Report the technical specifications of the MR scanner, including field strength, RF transmit coil type, maximal transmit power, and the number of head coil channels. Standard guidelines for proper reporting on MRI/fMRI parameters should be considered (Grainger, 2014; Nichols et al., 2017; Poldrack et al., 2008). |
| | | | 2.1.2. Report the details of MR conditionality that are demonstrated by the manufacturer of the tES equipment for specific conditions of use. |
| **2.2. tES-fMRI Setting Test - Safety Testing** | The safety of the tES-fMRI setting. | | 2.2.1. Report safety tests and respective details which include but are not limited to impedance testing, temperature testing (any temperature change under electrodes) and electric current tolerance testing, etc. with real human subjects or phantoms. Whenever the safety testing is referred to a previous study, it is still recommended to provide a brief description of the safety tests that have been considered. |
| | | | 2.2.2. Report the occurrence/absence of any safety incidents. |
| **2.3. tES-fMRI Setting Test - Subjective Intolerance Reporting** | The number of cases that have not tolerated the tES/fMRI session (even if it is zero). | | 2.3.1. Report the reasons that participants have not tolerated the tES/fMRI session if any (i.e., burning sensation, increased temperature, pain, shortness of breath, nausea, etc.). |
| **2.4. tES-fMRI Setting Test - Noise/Artifact** | The noise/artifact induced by the tES setting in the fMRI signal with real human subjects or phantoms before starting the study (It can be reported or referred to previous studies with the same setting). | | 2.4.1. Report or cite prior analysis on the degree to which the equipment alone, and the equipment during stimulation affects the SNR. Importantly, such analysis is specific to the protocol (electrode preparation, imaging sequence) such that claims cannot be automatically generalized without analysis. For instance, ~8% as described in (Antal et al., 2011) ("... SNR was hardly reduced with decreases ranging from 3 to 8% for the different ROIs and setups, even in the gray matter ROI in M1 targeted by tDCS....."). |
| | | | 2.4.2. Report how many participants, or runs were excluded from the analysis due to artifacts. Exclusion criteria should be reported as well (e.g., based on visual inspection or any data analysis tool that might detect artifacts for single runs). |
| | | | 2.4.3. Report the quantification of the possible increase in artifact or noise If the task-related fMRI requires the use of some other devices, such as tactile/pain stimulators, olfactory or juice machines, etc. (e.g., compare the noise/artifacts of the tES-fMRI setup alone with the tES-fMRI setup with the addition of the respective device). |
| | | | 2.4.4. Report baseline "pre-tES" fMRI as a part of the data acquisition sequence in the imaging session to investigate the effects/noise introduced by the tES setup per se (without any stimulation and within subject). Although this will not be sufficient to fully control for noise induced by tES administration with problems such as scanner drift, and the order effect. |
| | | | 2.4.5. Report any special fMRI processing measures or assessments that are used to deal with tES-induced imaging artifacts if applicable. |
| **2.5. Impedance Testing** | Impedance monitoring (i.e. before entering the scanner room and/or in the scanner room and/or inside the scanner and/or during scanning). | | 2.5.1. Report the impedance (i.e. cut off criterion programmed in the device, or measures on an individual basis with mean/range across groups before, during, and after scanning). |
| | | | 2.5.2. Report the methods applied to verify the current delivered inside the scanner (if any). Some devices already include an independent current meter and some investigators use their own external devices. |
| ***Methodological Factors*** | | | |
| **3.1. Concurrent tES-fMRI Timing** | The timing of concurrent tES within the fMRI paradigm. | | 3.1.1. Providing schematic diagrams is strongly encouraged to achieve maximum clarity for the reader. |
| | | | 3.1.2. Report carry-over effects between different stimulation conditions and different brain states. How such effects have been considered or mitigated should be discussed. |

| | | |
|---|---|---|
| **3.2. Imaging Session Timing** | The imaging events before and after concurrent tES-fMRI and respective sequences. | 3.2.1. Report the exact timing of all imaging events (structural or functional) before and after concurrent tES-fMRI. |
| | | 3.2.2. Report when the tES setup is placed on the participant e.g., if the tES setup was placed on the participant at the start of the tES-fMRI session (and was therefore on the participant during other non-fMRI sequences). |
| | | 3.2.3. In tACS studies, report how stimulation frequency is matched with TR. To reduce potential sources of biases in tACS-fMRI studies, the stimulation frequency should be set such that a full number of cycles fits into the TR of the functional measurement (Antal et al., 2014) (post-mortem study). Otherwise, the tissue polarization might be averaged over the time of one volume measured. |
| **3.3. tES Experience Report** | The assessment of the subjective experience of receiving tES inside the scanner. | 3.3.1. Report the general experience (comfort/fatigue) and participant's other experiences with the stimulation - as some tES montages/protocols might be more uncomfortable/perceptible than others when lying inside the scanner and this could be a confounder when comparing across stimulation montages. Options include: assessing participant ratings of symptoms for each condition, asking participants whether they perceived stimulation or not for each condition, reporting on the presence and intensity of phosphenes/tactile sensation (in the case of tACS), etc. This is important as it could show whether participants can differentiate between stimulation conditions (e.g., between active and sham stimulation, or between different frequencies (in the case of tACS). Having different side effects between sessions does not necessarily mean that subjects can discern and are unblinded. |
| | | 3.3.2. Report electric current tolerance for subject comfort (i.e. before entering scanner room (if technically possible) and/or in the scanner room and inside the scanner and/or during scanning (as appropriate)). |
| | | 3.3.3. Report any instructions, training, or exposure provided before the tES-fMRI session to make the experiment more convenient for the participants. |
| | | 3.3.4. Report the exact wording or provide citations of the questions or questionnaires used to report on the subjective experience of receiving tES inside the scanner in the article or its supplements. |
| **General Recommendations** | | 0.0.1. Report handedness of subject as a potential source of variability of tES-fMRI studies. This interaction could be addressed in relevant contexts either by limiting the sample to right-handed individuals, reporting handedness with quantitative standard instruments, or through methodological/analytical approaches which should be reported. |
| | | 0.0.2. If possible, present the online tES electrodes as additional bumps in the surface/mesh reconstruction. This is a good possibility to determine the exact location of the online electrodes. However, this non-biological reconstruction may also influence simulations, so performance of additional structural T1w and T2w scans without the electrodes whenever possible is advantageous. |

*We strongly recommend that this checklist be read in conjunction with the ContES checklist development and consensus paper. The paper should be cited when using the checklist as well.
